# Disentangling effects of ethnicity, deprivation, and payment source on obstetric outcomes in American primigravidae: A structural equation model of observational data

**DOI:** 10.1101/2024.10.30.24316452

**Authors:** Jonathan Williams

## Abstract

**Background:** Women from ethnic minorities have worse obstetric outcomes. Possible reasons for this are (1) social deprivation; (2) different standards of obstetric care; and (3) intrinsic ethnic differences. Here I aim to disentangle (1)-(3).

**Methods:** I constructed two path models of causal links between parental ethnicity and obstetric outcomes. The first, ‘no-racism’, model estimated independent causal effects of ethnicity, deprivation and payment source on pregnancy and birth outcomes. The second ‘realistic’ model additionally tested how far deprivation and payment source may mediate effects of ethnicity. Analyses of the models used Bayesian estimation. I analysed both the full sample of complete data and a random 1% sample.

**Findings:** Data were complete for 762786 births. The ‘no-racism’ model did not fit the data, but the ‘realistic’ model fitted adequately. It indicated that ethnicity, social deprivation, and private funding for care all adversely affected outcomes: (i) African American and Hispanic ethnicity caused deprivation; (ii) deprivation increased pregnancy hypertension, shortened gestation and reduced birthweight; (iii) private funding directly increased pregnancy hypertension and indirectly shortened gestation; (iv) participation in the Supplemental Nutrition Program for Women, Infants and Children (WIC) counteracted adverse effects of deprivation. (v) independently of (i)-(iv), ethnic-minority parents had shorter gestation and lighter babies.

**Interpretation:** Deprivation largely accounts for adverse obstetric outcomes in ethnic minorities. Private funding may also worsen pregnancy hypertension, but WIC improved outcomes. The uniformity of adverse birth outcomes for all ethnic minorities suggests that these result from a common factor, which may be systemic racism. Policies to reduce deprivation and increase government-funded care could importantly improve obstetric outcomes, irrespective of ethnicity.

**Funding:** none – I undertook the study at home.

**Research in Context:** *Evidence before this study:* Many studies during the past century have shown that ethnic minorities have worse social deprivation and worse access to health services. Ethnicity, deprivation and care can all determine health outcomes, and ethnic-minority mothers have worse obstetric outcomes. However, the independent contributions of ethnicity, deprivation and care to these adverse outcomes are unknown.

*Added value of this study:* I present here causal model of routine observational data that differentiates direct and indirect effects of ethnicity, deprivation and payment source on obstetric outcomes. The model allows (a) deprivation to mediate effects of ethnicity and (b) payment source to mediate effects of both ethnicity and deprivation. Hence, this model can disentangle the “intertwined” effects of ethnicity, deprivation and payment source on obstetric outcomes. The model also examines effects of participation in the Supplemental Nutrition Program for Women, Infants and Children on outcomes. The model fitted a 1% sample of the data after Bayesian estimation – so it bears interpretation as a representation of the real-world causal structure of the data. In the model, minority ethnicity causes deprivation and medical insurance and *all* of these factors independently determine adverse obstetric outcomes. Notably, medical insurance and private payment may increase the risk of pregnancy hypertension and consequently shorten gestation. Participation in WIC was beneficial.

*Implications of all the available evidence:* Causal modelling of routine natality data may allow effective audit of health care in its social context. Understanding causes of poor outcomes can enable prediction of effects of policy change. The present results indicate that policies to ameliorate social deprivation and expand access to WIC and government-subsidised care should improve obstetric outcomes – with long-term benefits for both mothers and their babies. Extrapolating beyond obstetrics, the present results may help to illuminate mechanisms of the healthcare crisis in America.

## Introduction

Social deprivation is a major determinant of health^1,2^ and ethnic minorities generally suffer more deprivation.^3,4^ In America, deprivation can determine the source of payment for healthcare,^5,6^ which can condition care and outcomes.^7–9^ This is because (a) deprived areas may have limited access to health services,^10,11^ and (b) doctors may not accept patients who lack medical insurance.^12,13^ Hence, social determinants of health are “deeply intertwined with … racial and rural disparities”.^14^

Obstetric outcomes are important, because they predict long-term health problems for both mothers and babies.^14–19^ Mothers from ethnic minorities have worse obstetric outcomes.^20–27^ These may reflect (1) direct effects of ethnicity;^20,28–31^ (2) social deprivation;^14,21,22^ or (3) differences in obstetric care between ethnic groups.^24,32–34^ It is important to disentangle (1)-(3), in order to inform both social policy and obstetric management.^14,35^ Previous studies separated effects of deprivation and ethnicity on obstetric outcomes^36–40^ but did not assess effects of payment source.^41–43^ Here, I aim to disentangle effects of all these “intertwined”^14^ variables on obstetric outcomes^36,44,45^ (see Fig 1), *via* causal modelling of routine observational data.^36,44–47^

**Figure 1.**
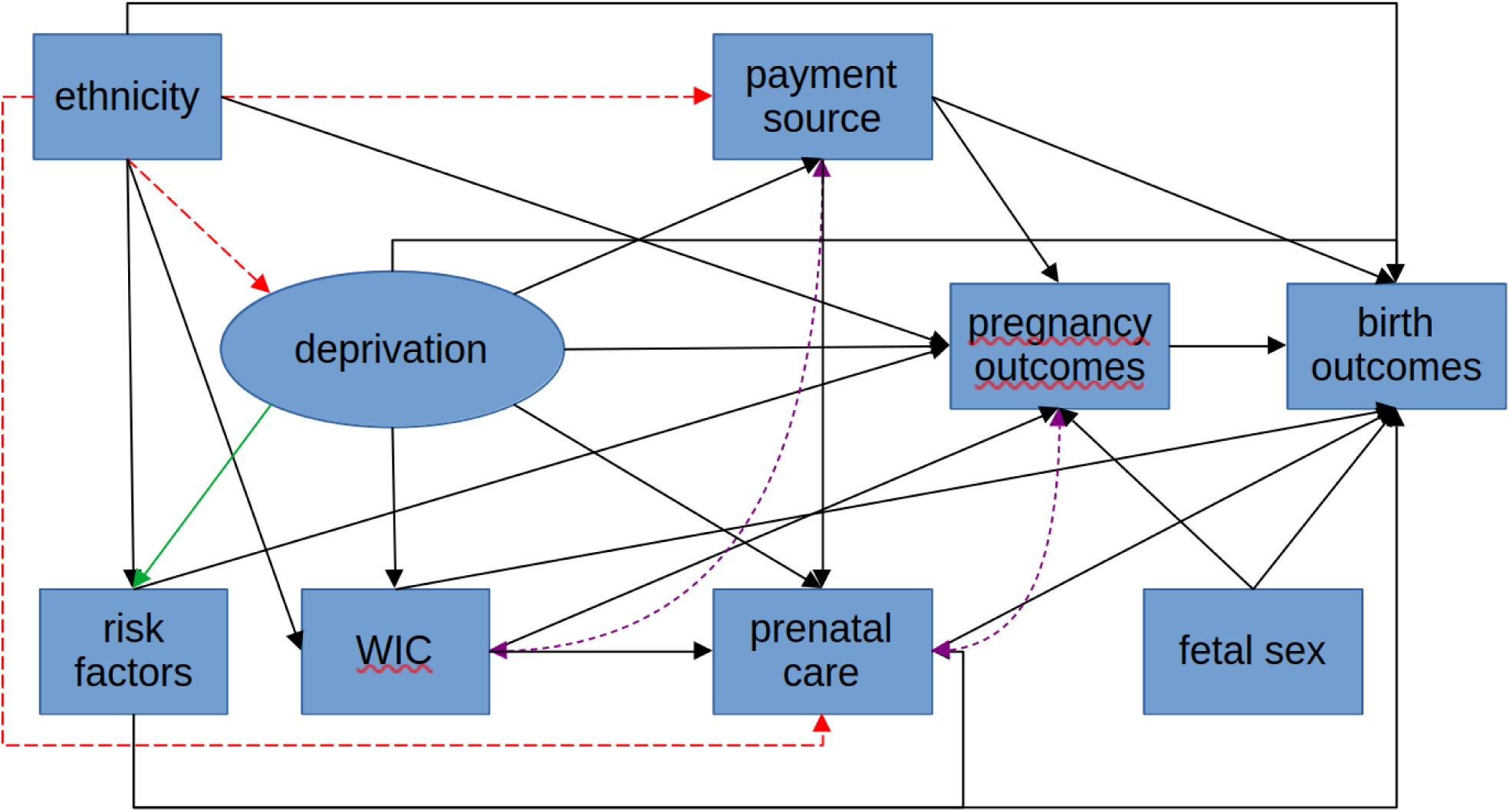
– a schematic diagram of the causal paths in the ‘realistic’ and ‘no-racism’ models. Legend: The diagram shows the causal paths and non-causal associations between ethnicity, deprivation, payment source, and other variables. Rectangular boxes represent manifest variables; the oval represents latent deprivation. Single-ended arrows represent causal paths, double-ended, mauve, dashed arrows represent correlations. The green arrow indicates that risk factors are manifest indicators of deprivation. The ‘no-racism’ model excluded the red dashed arrows. Supplementary Figure S1 shows more complete details of the study models’ path diagrams. Key: WIC = participation in the Supplemental Nutrition Program for Women, Infants and Children.

The temporal progression of pregnancy permits modelling plausible paths by which ethnicity, risk factors, deprivation and payment source may cause obstetric outcomes (Figure 1). This progression is relatively predictable, compared with many medical disorders. Hence, pregnancy and childbirth may provide a ‘unique opportunity’ as a model system to disentangle the “intertwined” effects of ethnicity, deprivation, and source of payment in both obstetric settings and for wider outcomes.^14^

Deprivation is a complex concept that has geographical, economic, social and health dimensions.^48^ Measures of material and social deprivation relate strongly to each other and to physical and mental health.^49^ Here, I assess deprivation using educational, social and health indices, including paternal educational level and age,^50^ together with maternal education, smoking, adult stature^51^ and BMI.^52,53^ I model ethnicity as a cause of deprivation and both deprivation and ethnicity as causes of risk factors and of the source of payment, prenatal care and outcomes (Figure 1). This model, therefore, embodies possible causal paths in the progression of pregnancy and birth.

Statistically, there are two approaches to test if deprivation, lack of medical insurance and prenatal care may mediate adverse effects of minority ethnicity on obstetric outcomes. The first is to assess the absolute fit of a ‘realistic’ model that includes these mediation effects. If this model fits the data adequately, then it may represent real-world social and clinical causes.^45,54,55^ A second approach is to test if the above ‘realistic’ model fits the data better than a ‘no-racism’ model that omits ethnicity’s effects on deprivation, source of payment and prenatal care. Here, I show that the ‘realistic’ model both outperforms the ‘no-racism’ model and can fit the data – which means that it may accurately represent real-world social and clinical causes of adverse obstetric outcomes.

## Methods

### Data and ethics

I analysed the US CDC’s natality data^56^ from primigravidae who delivered live infants in their third trimester during 2019. The CDC has described the data^57^ and Supplementary Table S1 shows the variables that I used. The data are anonymised and publicly-available,^56^ so no ethical approval is necessary.^58^ I use ‘ethnicity’ to include ‘race’, following the United Nations recommendation.^59^

### Data selection

I initially selected all primigravidae aged 14-45. I then excluded (i) births before the 35^th^ or after the 42^nd^ weeks of gestation; (ii) babies who weighed less than 1.5kg (which may be unhealthy^60^); (iii) births of babies with anencephaly (since these are non-viable); mothers with extreme values of (iv) height (<1.44m or >1.88m), or (v) pre-pregnancy BMI (<16.5kg/m^2^ or >40kg/m^2^), or (vi) pre-pregnancy hypertension or (vii) diabetes. Exclusions (iv)-(vii) may help to minimise pre-pregnancy maternal morbidities.^61^ Finally, I also excluded natality records with any missing data for the study variables (see Supplementary information for R code that generated the data-set).

### Data pre-processing

I coarsened measures of payment and ethnicity: (a) I dichotomised payment source into (i) government-aided (Medicaid, CHAMPUS/TRICARE, Indian Health Service or other governmental payment) and (ii) private payment (via private insurance, or self-payers);^5^ (b) I created a category of “Other” ethnicities for all Asian, American Indian/Alaskan (AIAN) or Native Hawaiian or Pacific Island (NHPI) parents, because there were few NHPI or AIAN parents and the “Asian” group is already diverse^56^ (the nHEA category is also genetically diverse^62^); (c) I dichotomised smoking as mothers who reported no smoking or who smoked at any time; (d) I pooled the lowest and 2^nd^-lowest categories of educational level (lowest = 1 = less than 8^th^ grade; 2^nd^-lowest = 2 = up to 12^th^ grade with no diploma) and the highest and 2^nd^-highest categories (highest = 8 = higher academic or professional degree; 2^nd^-highest = 7 = Master’s degree), for both mothers and fathers.

I re-scaled and centred parental ages, maternal BMI and height (see Supplementary Methods) to have mean ≈0 and variance ≈1. I centred initiation of pre-natal care on the 3^rd^ month of gestation and the duration of gestation on 39 weeks. I scaled birthweight to kg and centred it on 3.5kg.

### Model construction and estimation

I constructed a path model that estimated independent effects of ethnicity, deprivation and payment source on obstetric risk factors and outcomes (see Fig 1 and Fig S2). Note that the estimation fixes potential paths that are absent from the model at zero. Hence, the paths that are present can represent a causal model.^63,64^ Model estimation used Mplus.^65^ The Supplementary Information shows all programs and outputs.

The model defined deprivation as a reflective factor with six manifest indicators, that can tap long-term hardship^66,67^ (see Figure 2): (1) younger paternal age,^50,68,69^ (2-3) lower levels of paternal and maternal education^70,48,50^, (4) maternal smoking^71–74^, (5) shorter maternal stature^51,75–77^ and (6) higher pre-pregnancy maternal BMI.^71,53,78,79^. Minority ethnicity may evoke structural racism,^80–83^ that can strongly determine deprivation.^4^ However, deprivation occurs in all ethnic groups. Therefore, I aimed to derive estimates deprivation that are independent of ethnicity. To this end, I adjusted the manifest indicators of deprivation for ethnicity when estimating the latent deprivation factor.

**Figure 2:**
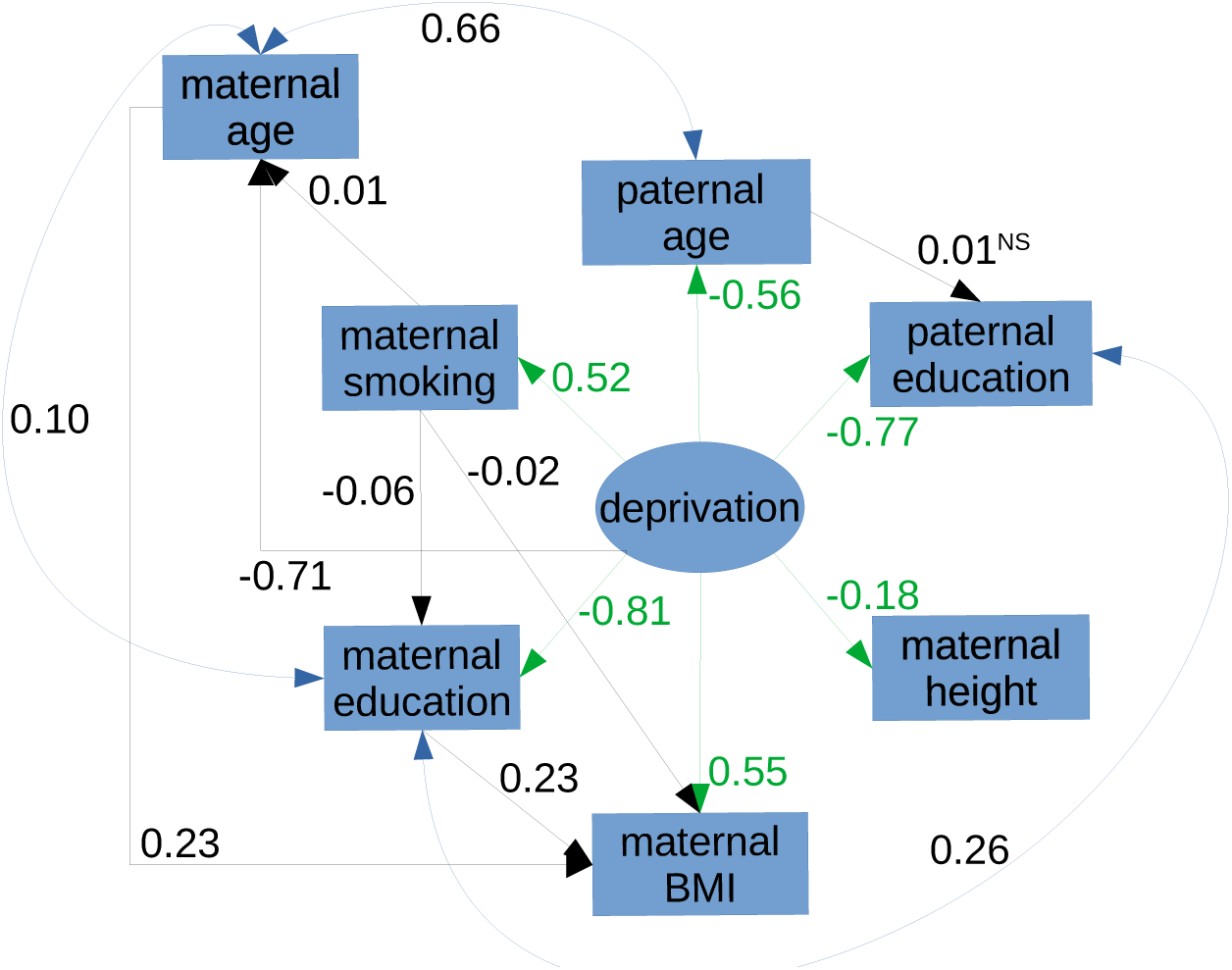
the structure of the deprivation factor. Legend: The inter-relations of parental characteristics and their loadings (red arrows) on the deprivation factor. Conventions for model symbols are the same as Figure 1. The values are the estimated standardised effects on each parental characteristic of increasing each putative causal factor by one notional standard deviation. Loadings of the latent variable “deprivation” are in red; single-ended black arrows show putative causal effects of maternal smoking, age and education on each other and on pre-pregnancy BMI; double-ended blue arrows show bidirectional correlations between maternal and paternal age and education and maternal smoking with paternal age. Abbreviation: NS = not significant (all other coefficients have p<0.001)

The 6 parental demographic factors, above, should show local independence,^84^ conditional on the deprivation factor. However, I additionally modelled causal effects and associations between parental demographic factors (see Figure 2). I assumed that (1) smoking can cause BMI^85,86^ and educational level (because smoking usually begins before education ends^87,88^); (2) longer maternal education may delay first pregnancy, but women may curtail education if they become pregnant – so the causal direction of the link between maternal age and education is uncertain; (3) paternal age can determine paternal education, but not vice versa; (4) assortative mating may strengthen correlations between parental ages and educational levels,^89–92^ over and above links due to ethnicity and deprivation; (5) deprivation precedes pregnancy – so, I modelled maternal age as an effect of deprivation; (6) private payment (private insurance or self-payers) and participation in WIC relate inversely, over-and-above their separate dependence on deprivation (Figures 2a-b); (7) BMI correlates directly with participation in WIC^93^ (in line with its direct correlation with deprivation^53,78,79^) and with private payment.^94^

The model’s deprivation factor does not use direct economic measures as manifest indicators. Participation in the Supplemental Food Program for Women, Infants and Children (WIC)is a direct indicator of material deprivation, because it requires families to have relatively low income.^95,96^ (although WIC may not index deprivation perfectly, because it may require participants to work^97^). Conversely, most people with medical insurance have paid work.^98^ Therefore, I tested the factor’s validity by assessing its effects on both WIC and on source of payment for obstetric care.

I tested if education may cause participation in WIC,^99^ over-and-above effects of ethnicity and deprivation. I allowed smoking status and BMI to correlate with both participation in WIC and source of payment. My rationale for including these correlations is that both smoking and BMI may increase participation in WIC and reduce insurance rates, by reducing disposable income.^71,78,100^ However, the models would not converge if I included smoking or BMI as *causes* of WIC and source of payment, but only converged if I included them as a *correlates*.

Finally, I compared the ‘realistic’ model with a ‘no-racism’ model that did not include effects of ethnicity on deprivation, source of payment or month of beginning pre-natal care (see Figure 1 and Figs. S2-S3). In effect, the ‘no-racism’ model allows ethnicity, deprivation and care (source of payment and initiation of prenatal care) to exert their effects separately, but not in combination. Hence, this comparison allows assessment of the “intertwined” effects of ethnicity with deprivation and care and so provides a pointer to the importance of racism in determining obstetric outcomes.

The model provided indirect effects from putative causal factors to outcomes. I present only those indirect effects that test if deprivation mediates adverse effects of non-nHEA ethnicity on outcomes. The Mplus outputs in the Supplementary Information contain complete tables of indirect effects.

The large sample size (∼¾ million) makes the analysis very sensitive, in two ways. (1) analyses of large samples can detect small effects of no practical importance. Therefore, I assessed significance of path coefficients in the Frequentist analysis of the full sample using the approximate Bayes Factor: |t|>sqrt(log(N)+10).^101^ I describe below only effects with |t|>4.86, which implies an approximate Bayes Factor (BF)>150. For the Bayesian estimation, I calculated approximate t-values from the Mplus output as: t=“Estimate” / “Posterior S.D.” and used the same criterion of |t| >4.86. This strategy parallels the “five-sigma” level of improbability that is the standard for statistical significance in physics.^102^ (2) Following from (1), the χ^2^ test is very sensitive for detecting lack of fit in large samples (>1000).^103^ Therefore, I assessed model fit in two ways: first, I used the Root Mean Square Error of Approximation (RMSEA), Comparative Fit Index (CFI), Tucker-Lewis Index (TLI) and Standardised Root Mean squared Residual (SRMR); I set RMSEA and SRMR values less than 0.025 as indicating acceptable fit;^104^ second, I analysed a random 1% sample (approximately 7800 births) that may be less sensitive to minor deviations from the model.

## Results

### Sample characteristics

The study sample comprised 762786 primigravidae with singleton live births in America in 2019 (see Table 1). The Supplementary Information shows the sample selection process (Figure S2) and differences between included and excluded mothers (Tables S2-S3).

**Table 1:** characteristics of the sample used for analysis, broken down by ethnic group.

| Variable | Ethnicity: | n-H European | African American | Hispanic | Other |
| --- | --- | --- | --- | --- | --- |
| Number of mothers |  | 451267 | 89356 | 136847 | 85316 |
| Number of fathers |  | 453111 | 104732 | 126795 | 78148 |
| Smoking mothers (%) |  | 6.8 | 3.0 | 1.7 | 1.7 |
| Maternal Education* |  | 6 (4 – 6) | 4 (3 – 6) | 4 (3 – 5) | 6 (4 – 7) |
| Paternal Education* |  | 5 (3 – 6) | 3 (3 – 5) | 3 (3 – 4) | 6 (4 – 7) |
| Maternal age (y) |  | 28 (24 – 31) | 24 (21 – 29) | 25 (21 – 29) | 30 (27 – 33) |
| Paternal age (y) |  | 30 (26 – 34) | 26 (22 – 31) | 27 (22 – 31) | 32 (29 – 35) |
| Maternal height (m) |  | 1.65 (1.60 – 1.70) | 1.63 (1.57 – 1.68) | 1.60 (1.55 – 1.65) | 1.60 (1.57 – 1.65) |
| Maternal BMI (kg/m<sup>2</sup>) |  | 24.4 (21.7 – 28.3) | 25.7 (22.2 – 30.1) | 25.3 (22.3 – 29.3) | 22.8 (20.5 – 25.8) |
| WIC participant (%) |  | 17.3 | 50.2 | 44.7 | 16.7 |
| Insured / self-payer (%) |  | 77.0 | 42.2 | 48.7 | 76.7 |
| Pre-natal care (month) |  | 2 (2 – 3) | 3 (2 – 4) | 3 (2 – 3) | 2 (2 – 3) |
| Pregnancy high BP (%) |  | 9.9 | 9.7 | 7.2 | 5.4 |
| Gestational diabetes (%) |  | 5.0 | 3.9 | 4.9 | 10.9 |
| Gestation (weeks) |  | 39 (38 – 40) | 39 (38 – 40) | 39 (38 – 40) | 39 (38 – 40) |
| Birthweight (kg) |  | 3.36 (3.06 – 3.66) | 3.18 (2.87 – 3.48) | 3.29 (3.01 – 3.58) | 3.20 (2.91 – 3.49) |
| Maternal deprivation score |  | -0.08 (-0.66 – 0.71) | 0.92 (0.23 – 1.43) | 0.93 (0.19 – 1.48) | -0.67 (-1.16 – 0.12) |
| Paternal deprivation score |  | -0.12 (-0.69 – 0.66) | 0.94 (0.24 – 1.45) | 0.95 (0.22 – 1.49) | -0.63 (-1.12 – 0.22) |
| Missing data (%) |  | 19.8 | 39.1 | 36.1 | 16.4 |

Key: n-H European =non-Hispanic European American; Cigs = cigarettes; high BP = hypertension; BMI = body mass index; WIC = participation in Supplementary Nutrition Program for Women, Infants and Children.

### Model fits

The ‘realistic’ model (Figures 1 and 2) fitted the data adequately, after WLSMV estimation (RMSEA = 0.015, 95% CI = 0.014-0.015; CFI = 0.999; TLI = 0.992; SRMR = 0.009), but not perfectly (χ^2^ = 5404, 33df, p<0.001). The ‘realistic’ model fitted equally imperfectly after Bayesian estimation (Posterior Predictive χ^2^ 95% CI = 3952 – 4212, 185 free parameters, p<0.001). The fit of the ‘no-racism’ model was much worse than that of the ‘realistic’ model ( Posterior Predictive χ^2^ 95% CI = 73936– 148737, 167 free parameters, p<0.001; Satorra-Bentler χ^2^ = 44771, 18df, p<0.001).

The ‘realistic’ model fitted a 1% random sample of the data (Posterior Predictive c^2^ 95% CI = −31.8 – 643.8, 185 free parameters, posterior predictive p=0.179; see Supplementary Information).

Since the ‘no-racism’ model performed poorly, I do not consider it further, here in the main report. Full details of the ‘no-racism’ model are available in the Supplementary Information, together with tables (S6a - S6c) comparing its path coefficients with those of the ‘realistic’ model.

### Structure of the deprivation factor

The latent dimension of deprivation loaded directly on maternal BMI and smoking, but inversely on paternal age and education, and on maternal education, age and height (Figure 2). Note that these loadings were independent of (i) effects of ethnic group on every indicator and on deprivation itself, (ii) correlations between paternal and maternal age and education, and (iii) effects of smoking on maternal education and BMI of smoking on BMI.

### Effects of ethnicity and deprivation on parental risk factors

Parental African American (AA) or Hispanic ethnicity increased deprivation (see Tables 1 & 3). In contrast, parents of Other ethnicities had less deprivation than non-Hispanic European American (nHEA) mothers (Tables 1 & 3a).

Deprivation worsened all parental risk factors (age, education, BMI and stature - Figure 2 and direct effects of Deprivation in Tables 1 & 2). Deprivation generally mediated apparent adverse effects of parental ethnicities on parental age, education and BMI. Hence, at equal levels of deprivation: (a) AA fathers were slightly older than nHEA fathers; (b) AA parents were, overall, better-educated than nHEA parents; (c) AA and Hispanic ethnicity did not increase maternal BMI; (d) AA and nHAE mothers were the same age and height (compare Tables 1 and 3). In contrast, direct effects of ethnicity predominantly determined the shorter stature of Hispanic and Other-ethnicity mothers and the lower self-reported smoking rates of most ethnicities (except AA fathers) (Table 2).

**Table 2:** Effects of ethnicity and deprivation on parental risk factors. Legend: values represent indirect, direct and total effects of parental minority ethnicities and deprivation on parental risk factors. Where possible, the table splits indirect causal effects into those mediated by deprivation (othD and dirD) or not (nonD): ‘othD’ means indirect effects that have multi-stage mediation via deprivation; ‘dirD’ means indirect effects for which deprivation is the sole mediator. Total effects sum the direct and any indirect effects that are present. Values in red are those where deprivation mediates effects of ethnicity; those in green are where direct effects of ethnicity predominate. Values in bold have p<0.001 and differ from zero by ≥5 posterior standard deviations. The first row shows estimates for non-smoking, non-Hispanic European American (nHEA) mothers with mean deprivation, age, education, BMI and height, who did not participate in WIC and received government-funded care. The nHEA values are standardised intercepts, with estimated means in the variable’s original scale (in brackets). All values on lower rows are deviations from the nHEA intercepts, expressed as 100*(standardised path coefficients) that estimate effects, in standard deviations, of a 1 standard deviation change in each cause. Notes: (a) direct and indirect effects may not sum to total effects, due to rounding error; (b) categorical cofficients use the logit link. Abbreviations: AFAMM/AFAMF = AA mother/father; MHISP/FHISP = Hispanic mother/father; OTHM/OTHF = Other-ethnicity mother/father; SOCDEP = deprivation.

a) deprivation, smoking, maternal height and BMI
| Cause\|Effect: | SOCDEP | Maternal smoking (CIG0) |  |  |  |  | Maternal height (MHT) |  |  |  |  | Maternal pre-pregnancy BMI (BMI) |  |  |  |  |
| --- | --- | --- | --- | --- | --- | --- | --- | --- | --- | --- | --- | --- | --- | --- | --- | --- |
| Type of cause | Direct | nonD | othD | dirD | direct | total | nonD | othD | dirD | direct | total | nonD | othD | dirD | direct | total |
| nHEA | 0 | **-145 (4.2%)** |  |  |  |  | **-5.0 (164.7cm)** |  |  |  |  | **-3.8 (24.8kg/m<sup>2</sup>)** |  |  |  |  |
| AFAMM | **6.1** | - | - | **3.2** | **-17.0** | **-13.9** | - | - | **-1.1** | 0.0 | **-1.1** | **0.6** | **-2.2** | **3.4** | 0.8 | **2.6** |
| MHISP | **13.5** | - | - | **7.0** | **-22.8** | **-15.8** | - | - | **-2.4** | **-18.6** | **-21.1** | 0.0 | **-4.9** | **7.5** | -0.6 | **1.9** |
| OTHM | **-14.5** | - | - | **-7.6** | **-8.5** | **-16.1** | - | - | **2.6** | **-21.5** | **-18.9** | **-0.8** | **5.3** | **-8.0** | **-8.3** | **-11.8** |
| AFAMF | **20.0** | - | - | **10.4** | **-6.1** | **4.3** | - | - | **-3.6** | - | **-3.6** | 0.2 | **-7.3** | **11.1** | - | **4.0** |
| FHISP | **19.1** | - | - | **9.9** | **-15.2** | **-5.3** | - | - | **-3.5** | - | **-3.5** | 0.4 | **-6.9** | **10.6** | - | **4.1** |
| OTHF | -0.5 | - | - | -0.2 | **-6.0** | **-6.3** | - | - | 0.1 | - | 0.1 | 0.2 | **0.2** | -0.3 | - | 0.1 |
| SOCDEP | - | - | - | - | **52.2** | **52.2** | - | - | - | **-18.1** | **-18.1** | - | - | - | **55.4** | **55.4** |
| MEDUC | - | - | - | - | - | - | - | - | - | - | - | - | - | - | **22.7** | **22.7** |
| Smoker/CIG0 | - | - | - | - | - | - | - | - | - | - | - | - | - | - | **-5.8** | **-5.8** |

b) parental age and education
| Cause\|Effect: | Maternal age (MAGE) |  |  |  |  | Paternal age (FAGE) |  |  |  |  | Maternal education (MEDUC) |  |  |  |  | Paternal education (FEDUC) |  |  |  |  |
| --- | --- | --- | --- | --- | --- | --- | --- | --- | --- | --- | --- | --- | --- | --- | --- | --- | --- | --- | --- | --- |
| Type of cause | nonD | othD | dirD | direct | total | nonD | othD | dirD | direct | total | nonD | othD | dirD | direct | total | nonD | othD | dirD | direct | total |
| nHEA | **54.4 (28.0 years)** |  |  |  |  | **2.2 (30.1 years)** |  |  |  |  | **-29.4 (70.2%)** |  |  |  |  | **-4.5 (52.9%)** |  |  |  |  |
| AFRAMM | -0.2 | 0 | **-4.3** | -0.1 | **-4.5** | - | - | **-3.4** | - | **-3.4** | **1.0** | -0.2 | **-4.9** | 0.6 | **-3.4** | - | 0 | **-4.6** | - | **-4.7** |
| MHISP | -0.3 | 0.1 | **-9.5** | **1.2** | **-8.5** | - | - | **-7.6** | - | **-7.6** | **1.3** | **-0.4** | **-10.9** | **-4.0** | **-13.9** | - | 0 | **-10.3** | - | **-10.4** |
| OTHM | -0.1 | -0.1 | **10.2** | -0.2 | **9.8** | - | - | **8.2** | - | **8.2** | **0.5** | **0.4** | **11.7** | **-3.9** | **8.8** | - | 0.1 | **11.1** | - | **11.2** |
| AFRAMF | -0.1 | 0.1 | **-14.1** | - | **-14.0** | - | - | **-11.2** | **1.5** | **-9.8** | **0.4** | **-0.6** | **-16.1** | - | **-16.4** | 0 | -0.1 | **-15.3** | **2.2** | **-13.2** |
| FHISP | -0.2 | 0.1 | **-13.4** | - | **-13.5** | - | - | **-10.7** | **0.3** | **-10.4** | **0.9** | **-0.6** | **-15.4** | - | **-15.1** | 0 | -0.1 | **-14.6** | **-4.2** | **-18.8** |
| OTHF | -0.1 | 0 | **0.3** | - | **0.3** | - | - | **0.3** | **2.7** | **3.0** | **0.4** | 0 | **0.4** | - | **0.7** | 0 | 0 | **0.4** | **2.6** | **2.9** |
| SOCDEP | - | - | - | **-70.6** | **-70.6** | - | - | - | **-56.3** | **-56.3** | - | - | - | **-80.6** | **-80.6** | - | - | - | **-76.5** | **-76.5** |

**Table 3:** Effects of ethnicity, deprivation, and parental characteristics on payment source, participation in WIC and initiation of pre-natal care. Legend: values represent indirect, direct and total effects of maternal and paternal minority ethnicities on deprivation, participation in WIC, source of payment and initiation of prenatal care. The model’s structure, labels, values, and conventions are similar to those of Table 3. Note: ‘nonD’, ‘othD’, (dirD’ and ‘direct’ effects may not sum exactly to the ‘Total’effects, due to non-linearity of the logit link (for WIC and payment source) or rounding error (for start of pre-natal care). Cells with grey backgrounds are not directly relevant to the task of disentangling effects of deprivation, ethnicity and sources of funding. However, these effects may differ between nHEA and other ethnicities - see Tables S7a-l in the Supplementary Information. Abbreviations as in Table 2, plus FAGE/MAGE = paternal/maternal age; FEDUC/MEDUC = paternal/maternal education; CIG0 = maternal smoking history (0/1)

| Cause \| Effect: | Participation in WIC (WIC) |  |  |  |  | Insured or self-payer (PAYER) |  |  |  |  | Month began prenatal care (PRECAREM) |  |  |  |  |
| --- | --- | --- | --- | --- | --- | --- | --- | --- | --- | --- | --- | --- | --- | --- | --- |
| Type of cause | nonD | othD | dirD | direct | total | nonD | othD | dirD | direct | total | nonD | othD | dirD | direct | total |
| nHEA | **-94.0** (8.1%) |  |  |  |  | **73.8** (87.9%) |  |  |  |  | **7.3** (2.71 months) |  |  |  |  |
| AFRAMM | 0 | 0.1 | **4.2** | **7.1** | **11.5** | - | **0.3** | **-4.8** | **-6.8** | **-11.3** | -0.1 | **0.7** | 0.1 | **3.6** | **4.5** |
| MHISP | -0.2 | 0.3 | **9.4** | **5.4** | **14.9** | - | **0.7** | **-10.8** | **-4.6** | **-14.6** | -0.3 | **1.7** | 0.3 | **1.7** | **3.6** |
| OTHM | -0.1 | -0.2 | **-10.2** | **7.0** | **-3.5** | - | **-0.7** | **11.6** | **-9.2** | **1.7** | **0.9** | **-1.7** | -0.3 | **2.5** | **1.2** |
| AFRAMF | 0 | 0.3 | **14.0** | **4.7** | **19.0** | -0.1 | **1.0** | **-16.0** | **-3.2** | **-18.3** | 0 | **2.7** | 0.4 | **2.2** | **5.2** |
| FHISP | 0 | 0.3 | **13.3** | **4.8** | **18.5** | 0.3 | **1.0** | **-15.2** | **-2.7** | **-16.6** | -0.4 | **2.4** | 0.4 | **1.1** | **3.6** |
| OTHF | 0 | 0 | -0.3 | **4.7** | **4.4** | -0.2 | 0 | 0.4 | **-3.1** | **-3.0** | 0 | 0 | 0 | **1.5** | **1.4** |
| SOCDEP | - | **1.5** | - | **70.0** | **71.4** | - | **5.1** | - | **-80.0** | **-74.8** | - | **13.4** | - | **2.0** | **15.3** |
| FAGE | - | - | - | - | - | 0 | - | - | **-0.2** | **-0.3** | 0 | - | - | - | 0 |
| MAGE | - | - | - | **-5.9** | **-5.9** | - | - | - | - | - | 0 | - | - | **-3.3** | **-3.4** |
| FEDUC | - | - | - | - | - | - | - | - | **-6.6** | **-6.6** | **0.9** | - | - | - | **0.9** |
| MEDUC | - | - | - | **3.2** | **3.2** | - | - | - | - | - | **-0.6** | - | - | **-5.9** | **-6.5** |
| CIG0 | **-0.3** | - | - | - | **-0.3** | - | - | - | - | - | 0.3 | - | - | **2.4** | **2.8** |
| BMI | - | - | - | - | - | - | - | - | - | - | - | - | - | **-1.9** | **-1.9** |
| WIC | - | - | - | - | - | - | - | - | - | - | - | - | - | **-6.6** | **-6.6** |
| PAYER | - | - | - | - | - | - | - | - | - | - | - | - | - | **-13.3** | **-13.3** |

One-stage and multi-stage indirect causal effects involving deprivation were often different. For example, single- and multi-stage indirect effects of deprivation mediated opposite effects on BMI in ethnic minority mothers (Table 2a). Most indirect effects involved deprivation – most of those that did not involve deprivation were small beneficial effects on maternal education (Table 2a).

### Effects of ethnicity on deprivation and source of payment and WIC participation

Deprivation increased participation in WIC and reduced rates of private payment (Table 3). Independently, mothers and fathers of all minority ethnicities were less likely to pay privately (via insurance or directly) and more likely to participate in WIC (Tables 1 and 3).

### Effects of ethnicity, deprivation, payment source and WIC on gestational diabetes and hypertension

Different maternal ethnicities showed different patterns of direct effects on gestational diabetes (PRGDM) and pregnancy hypertension (PRGBP) (Tables 1 and 4): (a) both maternal and paternal Hispanic ethnicity directly reduced PRGBP; (b) both maternal and paternal Other ethnicity directly reduced PRGBP, but increased PRGDM – the direct effect of maternal Other ethnicity was almost three times as strong as that of Deprivation; and (c) AA paternity directly reduced PRGDM (all compared with nHEA fathers). Paternal AA and Hispanic ethnicity also increased PRGBP, indirectly – mainly via deprivation (Table 4).

**Table 4:**
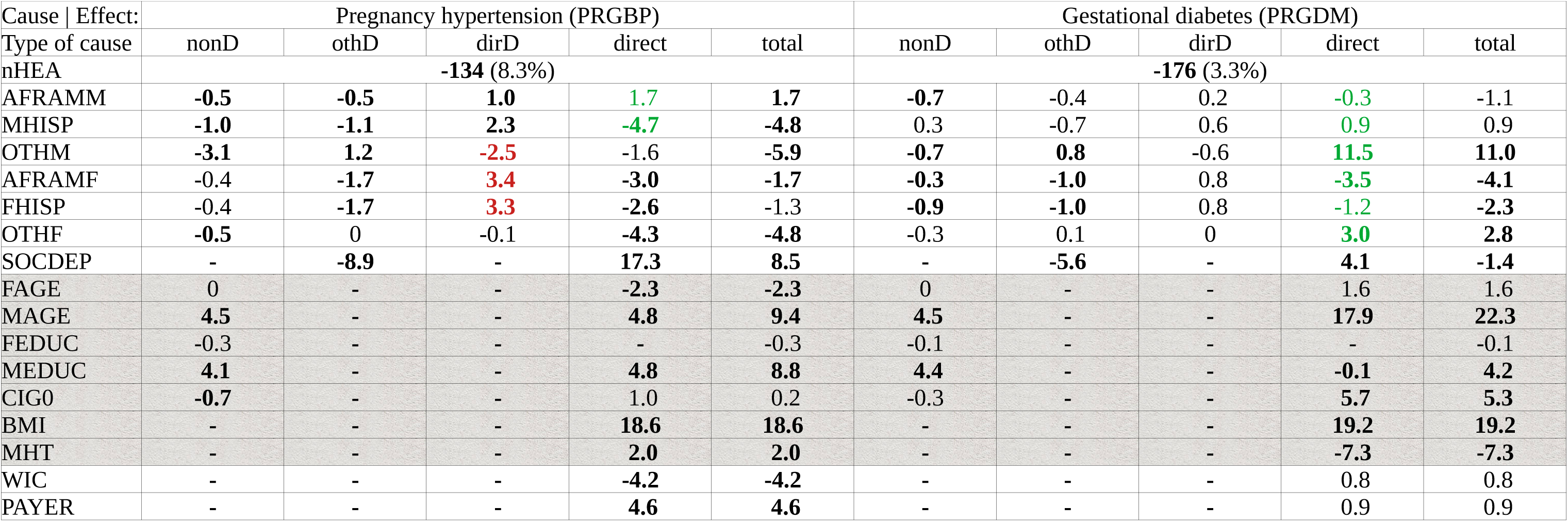
Indirect and total effects of all the model’s potential causal variables on pregnancy hypertension and gestational diabetes. Legend: values represent indirect, direct and total effects of all potentially-causal factors on participation on the proportions of mothers with pregnancy hypertension (PRGBP) or gestational diabetes (PRGDM). The table’s structure, labels, values, and conventions are similar to those of Table 3. Coefficients for deprivation represent the standardized change of the outcome per standard deviation change of the putative causal variable. Note: ‘nonD’, ‘othD’, ‘dirD’ and ‘direct’ effects may not sum exactly to the ‘Total’ effects, due to nonlinearity of the logit link (for PRGBP and PRGDM) or rounding error (for gestation and birthweight). Abbreviations as Tables 3 and 4. Cells with grey backgrounds are not directly relevant to the task of disentangling effects of deprivation, ethnicity and sources of funding. However, these effects may differ between nHEA and other ethnicities - see Tables S7a-l in the Supplementary Information.

Deprivation *per se* directly increased PRGBP, but its indirect effects reduced PRGBP and so partly cancelled its direct effect. This reflects deprivation’s loadings on younger maternal age, lower maternal education and shorter maternal height – all of which directly reduce PRGBP in the model. Private payment directly increased PRGBP, but participation in WIC reduced it (Table 5).

**Table 5:** Indirect and total effects of all the model’s potential causal variables on duration of gestation and birthweight. Legend: values represent indirect, direct and total effects of all potentially-causal factors on participation on the duration of gestation (OBGEST) and birthweight (BWT). The table’s structure, labels, values, and conventions are similar to those of Table 3. Coefficients for deprivation represent the standardized change of the outcome per standard deviation change of the putative causal variable. Note: ‘nonD’, ‘othD’, ‘dirD’ and ‘direct’ effects may not sum exactly to the ‘Total’ effects, due to rounding error. Abbreviations as Tables 3 and 4. Cells with grey backgrounds are not directly relevant to the task of disentangling effects of deprivation, ethnicity and sources of funding. However, these effects may differ between nHEA and other ethnicities - see Tables S7a-l in the Supplementary Information.

| Cause \| Effect: | Duration of gestation (OBGEST) |  |  |  |  | Birthweight (BWT) |  |  |  |  |
| --- | --- | --- | --- | --- | --- | --- | --- | --- | --- | --- |
| Type of cause | nonD | othD | dirD | direct | total | nonD | othD | dirD | direct | total |
| nHEA | **7.6 (39.1 weeks)** |  |  |  |  | **-40.3 (3.314kg)** |  |  |  |  |
| AFAMM | **0.4** | **0.5** | **-0.5** | **-2.1** | **-2.2** | **1.3** | -0.1 | -0.4 | **-9.8** | **-8.9** |
| MHISP | **1.7** | 0.3 | **-1.2** | **-1.4** | -0.9 | **-0.8** | 0.2 | **-0.9** | -0.4 | **-2.0** |
| OTHM | **-1.0** | -0.4 | **1.3** | **-3.2** | **-3.3** | **-4.0** | -0.1 | **1.0** | **-2.6** | **-5.7** |
| AFAMF | **1.9** | **0.5** | **-1.8** | **-2.7** | **-2.1** | **1.1** | 0.1 | **-1.4** | **-4.4** | **-4.5** |
| FHISP | **1.4** | **0.5** | **-1.7** | **-1.6** | **-1.4** | **1.7** | 0.1 | **-1.3** | **-4.2** | **-3.6** |
| OTHF | **1.4** | 0 | 0 | **-0.7** | **0.6** | **1.3** | 0 | 0 | **-7.9** | **-6.5** |
| SOCDEP | - | **2.7** | - | **-9** | **-6.2** | - | 0.6 | - | **-6.9** | **-6.2** |
| FAGE | **0.5** | - | - | -0.1 | 0.4 | 0.4 | - | - | **-1.5** | **-1.1** |
| MAGE | **-4.7** | - | - | 0.4 | **-4.2** | **1.6** | - | - | **-3.6** | **-2.0** |
| FEDUC | 0.2 | - | - | - | 0.3 | 0.1 | - | - | - | 0.1 |
| MEDUC | **-1.9** | - | - | -2.0 | **-3.9** | **1.7** | - | - | **-2.9** | **-1.0** |
| CIG0 | **-0.8** | - | - | **-0.2** | **-1.0** | -0.4 | - | - | **-7.3** | **-7.7** |
| BMI | **-8.8** | - | - | **7.8** | -0.9 | **-3.6** | - | - | **15.6** | **12.0** |
| MHT | 0.5 | - | - | **3.4** | **3.9** | -0.3 | - | - | **19.3** | **19.1** |
| WIC | 0.9 | - | - | 1.2 | **2.1** | **0.8** | - | - | 1.2 | **2.0** |
| PAYER | **-2.1** | - | - | -1.8 | **-4.0** | **-0.8** | - | - | **0** | **-0.8** |
| PRECAREM | - | - | - | **4.9** | **4.9** | 0 | - | - | -0.1 | -0.1 |
| PRGBP | - | - | - | **-31.5** | **-31.5** | - | - | - | **-18.5** | **-18.5** |
| PRGDM | - | - | - | **-14.4** | **-14.4** | - | - | - | **-1.2** | **-1.2** |

### Effects of ethnicity, deprivation, private payment and WIC on birthweight and duration of gestation

Deprivation directly reduced both birthweight and the duration of gestation (Table 5). Additionally, *all* minority ethnicities directly worsened these birth outcomes, compared with nHEA mothers (Table 5). Private payment indirectly shortened gestation, via PRGBP and earlier prenatal care (see Supplementary Information), but slightly increased birthweight (Table 5). Participation in WIC lengthened gestation both directly and indirectly, and also increased birthweight (Table 5).

### Indirect effects of maternal ethnicity and deprivation on outcomes

The model estimated 164 indirect causal paths from each maternal ethnicity to birthweight and duration of gestation. Many indirect effects were antagonistic (for example, AA ethnicity reduced duration of gestation via deprivation, but increased it via WIC), so that they partly cancelled each other. There are too many indirect effects to describe in detail and most were very small. The Supplementary Information shows all indirect effects from ethnicity, demographic factors, source of payment and and deprivation to obstetric outcomes. I summarise only a few illustrative indirect effects here and in the Supplementary Information.

The overall indirect effects of ethnicities on outcomes were inconsistent (see Table 5 and Mplus outputs in the Supplementary Information). Notably, (i) the sum of *indirect* effects of maternal AA ethnicity on birthweight was slightly positive (Table 5); and (ii) both maternal and paternal AA ethnicity showed only small *indirect* adverse effects on lower birthweight via deprivation. Deprivation’s *indirect* effects included lower risks of PRGBP (mainly via WIC) and PRGDM (mainly via lower maternal age), but slightly longer gestation (mainly via WIC) (see Table 3 and Mplus outputs). Private payment *indirectly* reduced the duration of gestation – mainly via higher rates of PRGBP and earlier initiation of pre-natal care (Table 5).

### Heterogeneity between ethnicities

The model here shows only main effects of ethnicity on different demographic and outcome variables. I assessed heterogeneity in effects of ethnicity by comparing separate analyses of each ethnic group – see Supplementary information.

## Discussion

The ‘realistic’ model aimed to disentangle effects of deprivation, ethnicity and payment source on obstetric risks and outcomes. This model indicates that: (1) Deprivation is a major cause of obstetric risks and adverse outcomes, independently of ethnicity. (2) Deprivation may mediate many apparent adverse effects of African American (AA) or Hispanic ethnicity on obstetric risks. (3) Private funding of obstetric care may cause pregnancy hypertension (PRGBP) and so reduce the length of gestation and birthweight (4) Participation in WIC can ameliorate deprivation’s effects. (5) AA, Hispanic and Other ethnicities *all* shorten gestation and lower birthweight directly (independently of deprivation and maternal risk factors). These results fit with extensive evidence that deprivation and ethnicity are major causes of morbidity and mortality.^1,105–108,82^ Hence, policies to reduce deprivation and increase public funding of obstetric services and participation in WIC may improve outcomes for most mothers and babies.

### Model validity

The ‘realistic’ model fitted the 1% sample of the data adequately. This indicates that the model can represent the causal structure of the real-world data-generating processes.^45,54,55^ Additionally, the model’s deprivation factor had predictive validity in that it directly determined (a) two economic indicators (payment source and participation in WIC), (b) a maternal risk factor (maternal age), and (c) adverse obstetric outcomes (PRGBP, shorter gestation and lower birthweight) – all of which associate with established measures of deprivation.^109–112^ Further, strengthening the factor’s construct validity, smoking and higher BMI can worsen income and housing, as well as worsening health.^71,78^ To test the validity of the ‘realistic’ model more directly, I constructed a second (’no-racism’) model that omitted ethnicity’s causal effects on deprivation and indicators of obstetric care (see Figure 1 and Supplementary Information). This ‘no-racism’ model did not fit the data. Finally, the present findings are consistent with a previous path model of deprivation’s effects,^36^ which indicates they are reliable. Further studies should test how the present deprivation factor relates to other measures of personal and neighbourhood disadvantage.^113–115^

### Model interpretation

The ‘realistic’ model uses parental risk factors as indicators of deprivation, but also allows ethnicity to determine both the risk factors and deprivation (see Methods). Hence it estimates both direct and indirect effects of ethnicity and deprivation on outcomes. The model adjusts direct effects (Tables 3-5) for preceding effects, so that each is interpretable as an independent cause. Overall, in the model, adjustment for deprivation ameliorates or even *reverses* apparent risk factors in ethnic minority parents. I discuss this further, below and in the Supplementary Information.

The ‘realistic’ model aimed to extract causal effects from correlations between ethnicity, risk factors, deprivation, funding sources and obstetric outcomes. Two examples of this are: (1) smoking correlates with young maternal age in both the raw data and the ‘no-racism’ model (see Supplementary Tables); in contrast the results of the ‘realistic’ model align with experimental evidence that smoking directly reduces fecundity;^116,117^ (2) The ‘realistic’ model showed that smoking directly lowered birthweight, independently of deprivation and other risk factors– in line with causal evidence that tobacco use can directly reduce birthweight,^118^ The consistency between the ‘realistic’ model and external causal studies indicate that it can disentangle “intertwined”^14^ causal effects of ethnicity, deprivation, maternal risk factors and payment source on outcomes.

Deprivation showed large effects on risk factors, payment source and participation in WIC. In contrast, after accounting for deprivation, most causal direct effects on obstetric outcomes were small and the model explained small proportions of the variance (*R*^2^) of the main outcomes (Table 2). Götz and colleagues have argued that small effect sizes are likely in real-world situations, where many uncontrolled variables affect each outcome.^119^ Here, after accounting for deprivation, ethnicity and payment source, known risk factors may be relatively unimportant for outcomes, compared with unmeasured factors (e.g. lifestyle choices, or environmental pollutants^120^).

### Smoking

Smoking is a major obstetric risk factor.^121,122^ Here, parents of ethnic minorities self-reported low rates of maternal smoking (Tables 1 and 2). This is consistent with previous obstetric data (Table 1 in^61,123,124^), but contrasts with objective evidence and self-reports in other settings that ethnic minorities smoke as much as non-Hispanic European Americans.^125–127^ The present results indicate that smoking in AA and Hispanic mothers results partly from deprivation. The contrast with non-obstetric settings implies that ethnic minority mothers may under-report their smoking in obstetric settings. Such under-reporting may reflect miscommunication or cultural distrust of obstetric clinicians.^108,128,129^ Under-reporting would have important consequences, here, because (a) the model uses self-reported smoking as a manifest indicator of deprivation and (b) smoking reduces birthweight.^118^ So, if ethnic minorities under-report smoking, then the model may under-estimate both their deprivation and adverse effects of both deprivation and smoking. Further studies should test objectively if primigravidae of minority ethnicities smoke less before and during pregnancy.

### Effects of ethnicity on deprivation, source of payment and initiation of prenatal care

AA and Hispanic parental ethnicity directly increased deprivation, but maternal Other ethnicities reduced it. Note that the model adjusted its manifest indicators of deprivation for parental ethnicity (e.g., shorter maternal stature loaded on deprivation even though both Hispanic and Other-ethnicity mothers were shorter than nHEA mothers). Hence, the deprivation factor reflects covariation between its indicators that is common to all ethnicities, including nHEA mothers.

All ethnic-minority parents were less likely than nHEA parents to pay for obstetric care via private insurance, or directly. This difference was apparent in the raw data and persisted after adjustment for deprivation (compare Tables 1 and 3). The uniformity of the effect – for all ethnic-minority parents – implies that it reflects a common cause, e.g. systemic racism, or income inequality.^130–132^

All ethnic-minority parents delayed initiation of prenatal care, in line with earlier reports.^133–135^ This difference was apparent in the raw data and persisted after adjustment for deprivation (compare Tables 1 and 3). Again, the uniformity of the effect – for all ethnic-minority parents – implies that it reflects a common cause, e.g. systemic racism, or income inequality.^130–132^

### Effects of ethnicity on outcomes

Minority ethnicities showed different direct effects on pregnancy problems – PRGBP and gestational diabetes (PRGDM). In particular, ‘Other’ (mostly, Asian) ethnicities developed more PRGDM, but Hispanic parentage reduced rates of PRGBP..^136^ This may reflect ethnicity-specific causes, such as cultural practices (e.g. diet^137,138^), or genetic heritage.^30,139,81,140^

In contrast to the ethnicity-specific effects on PRGBP and PRGDM, all minority ethnicities *directly* caused shorter gestation and lower birthweight. These results are in line with previous findings.^28,141,142^ Lower birthweights may partly reflect genetic factors in AA and Asian mothers;^30^ but babies born to fathers of all ethnic minorities have lower birthweight babies in the present study,^31^ and genetics do not completely account for lower birthweight in AA and Asian babies.^30,143^ The uniformity – for all minority ethnicities – of these direct effects on birth outcomes implies that they may partly reflect a common cause, such as vitamin D deficiency^144^ or systemic racism.^4,145–149,131,150–152,143^ Further studies should test these possibilities.

### Effects of deprivation on parental risk factors

Deprivation caused markedly worse obstetric risk factors for nHEA mothers in the present study, in line with previous reports (see Introduction & Table 2). Deprivation also mediated apparent adverse effects of ethnicity on risk factors in the raw data. For example, AA fathers were younger than nHEA fathers and AA parents, overall, received less education than nHEA parents (see Table 1) – but after adjustment for deprivation AA parents showed the opposite pattern of direct effects (see Table 2). These results are broadly consistent with previous findings that controlling for deprivation can ameliorate apparent adverse effects of ethnicity.^153,154^ However, ethnicity explains only 1/6th of the variation in deprivation – so policies to reduce deprivation would help mostly nHEA mothers.

### Effects of deprivation and ethnicity on source of payment and participation in WIC

AA and Hispanic parents were less likely to pay privately (Table 1) and Deprivation strongly reduced private payment (Table 3). Moreover, after accounting for deprivation, parents of *all* minority ethnicities were less likely to pay privately. Deprivation’s effect on government payment supports the validity of the present deprivation factor (see above). Perhaps the most likely reason for residual low rates of private payment by ethnic-minority parents is education-adjusted income inequalities and/or unemployment,^132^ because ethnic minorities are more likely to lose private medical insurance during their reproductive years.^155^ Further studies should test this possibility.

Deprivation strongly increased participation in WIC. Moreover, even after accounting for deprivation, parents of minority ethnicities directly increased participation in WIC (in line with previous findings^99^). Strikingly, higher levels of maternal education predicted participation in WIC (see Table 3), which contrasts with previous findings that WIC participants had lower levels of education *even after* adjusting for income.^99^ A possible explanation of this contrast is that the present model accounts more accurately for causes of WIC via deprivation and ethnicity. In this case, the present findings indicate that lower educational level may present a barrier to participation in WIC, among women who would otherwise be eligible. Further studies should test this possibility.

### Effects of deprivation on outcomes

Deprivation directly increased the risk of PRGBP, shortened gestation and reduced birthweight. These effects are consistent with previous results.^156,157,25^ However, the present findings clarify that deprivation can adversely affect outcomes independently of ethnicity, payment source, smoking, etc. (see Figs. 1-2). Further studies should clarify the mechanisms of these adverse effects.

### Effects of private payment on outcomes

Private payment directly determined higher rates of PRGBP. This fits with independent data (rows 1-2 of Insurance *x* Severe Preecplampsia in Table 1 in^58^: χ^2^ = 240, 1df, p<0.001; insurance *x* any hypertension in Table 1 in^158^: χ^2^=35.5, 1df, p<0.001), strengthening its reliability. Possible explanations for this are: (1) Financial stresses of funding medical care could directly cause PRGBP;^cf159,160^ or (2) clinicians have lower thresholds for diagnosing PRGBP in women who pay privately.^161–163^ This second possibility could also explain why women with Medicaid funding have worse PRGBP and more preeclampsia.^164^ It also fits the view that over-treatment may be common in America,^165^ particularly in the nHEA population,^166^ which has higher rates of private insurance coverage.^167^ PRGBP can cause adverse sequelae,^168,169^ including shorter gestation (due to pre-term induction) and lower birthweight. So, the present result mandates further studies to test (1) and (2).

### Effects of participation in WIC on outcomes

Participation in WIC directly lowered rates of PRGBP and lengthened gestation (see Table 4). These effects fit previous findings that WIC increased birthweight^170^ and reduced maternal mortality.^171^ Participation in WIC may be indicative of deprivation that is generally detrimental for maternal health, but – after accounting for deprivation – WIC could prevent PRGBP by reducing financial stresses.^96^ WIC’s beneficial effects contrast with the adverse effect of deprivation (see above), which highlights the potential to improve outcomes *via* policies that reduce deprivation.^cf159,172^

### Effects of pre-natal care on outcomes

All ethnic minority parents and smokers delayed starting pre-natal care (PNC).^see^ ^also108^ Every other maternal factor caused earlier PNC. However, starting PNC earlier determined with slightly higher risk of PRGBP, and slightly shorter gestation. The inverse correlation with PRGBP may occur because symptoms of early-onset PRGBP prompt some women to seek PNC. But, against this, starting PNC later did not also correlate with lower risk of PRGDM (which could also cause symptoms). An alternative explanation is that maternal anxiety about pregnancy caused earlier entry to PNC, increased PRGBP and shortened gestation.^cf159,173^ Further studies should test this possibility.

### Strengths of the study

The present study’s strengths are: (1) it used routine electronic birth certification (EBC) data from a large national sample of primigravidae; (2) the model’s causal structure provides an “open theory”,^63^ which is a step towards explanation and prediction;^54,63,64,55,45^ (3) the adequacy of the ‘realistic’ model’s fit implies that its structure can reflect clinical realities; (4) the analysis quantifies the model’s imperfect fit; (5) by excluding mothers and babies with extreme physical characteristics, the present results may more accurately describe the great majority of births.

The ‘realistic’ model simply represents plausible clinical causal pathways. Consequently, it may be non-identified^174^ and it does not try to avoid important biases that can cloud causal interpretation of observational data^174–176^ – such as “butterfly bias”, “collider bias” or “recanting witnesses”. Even so, most of the present findings (a) fit with previous reports, (b) parallel external causal analyses that used interventional methods (where available), and (c) appear plausible (*contra* known instances of collider or butterfly bias – e.g.^45,177–179^). Moreover, the ‘realistic’ model fits a 1% sample of the data. Therefore, the model may represent the real-world causal structure of the data-generating processes,^45,54,55^ which provides a basis for designing interventional studies to test its predictions.

### Limitations of the study

The study had important limitations: (1) Its sample excluded 1/3 of American primigravidae (see Methods), who overall had both worse deprivation and worse outcomes (see Supplementary Tables S2-S4). The main reason for exclusion was missing paternal ethnicity, which was more common for mothers of ethnic minorities. Given the concordance between parental ethnicities (Supplementary Table S5), missing ethnicity was probably more likely for AA and Hispanic fathers. Therefore, this selection bias almost certainly caused the model to *under*-estimate adverse impacts of deprivation and ethnicity.^180^ I discuss this further in the Supplementary Information. (2) The proportion of missing data may also vary geographically,^181^ so geopolitical factors may partly obscure effects of deprivation and/or ethnicity. Further studies should include geopolitical factors in the present model, in order to disentangle their effects on outcomes from those of deprivation, ethnicity and source of payment for care. (3) Electronic Birth Certificate data can be inaccurate^181,182^ - particularly for diagnoses of PRGBP and classification of self-payers.^181,182^ Again, it is likely that such inaccuracies would lead to *under-*estimation of adverse impacts of deprivation and ethnicity. (4) The model only includes the initiation of pre-natal care and not its frequency or quality. (5) The model does not account for obstetric interventions (e.g. early delivery in PRGBP) that may moderate effects of risk factors on outcomes. However, such interventions are links in causal chains that have earlier origins, and it is reasonable to view final birth outcomes as results of those earlier causes. Nevertheless, (2)-(5) may distort the model’s estimates of causal effects.

### Model fit

The ‘no-racism’ model, that excluded deprivation, did not fit the data, but the ‘realistic’ model fitted the data adequately, but imperfectly. The imperfect fit may be due to the large sample size, because this makes the analysis very sensitive to detect not only causal paths that are present in the model, but also small unknown causes that are absent from the model. In line with this explanation, estimation of the model using a 1% sample generated the same pattern of results and posterior predictive probability >0.10, signifying an adequate, though approximate, fit.

The model’s imperfect fit to the data of the full sample may reflect (a) remediable limitations of the present methods and/or (b) substantive deficiencies of the model. The most obvious remediable limitation is coarsening of education, smoking and ethnicity data – especially pooling data from Alaskan and American Indian mothers with those of American and Native Pacific Islanders and Asians, since the former two groups generally experience more social deprivation,^183^ while the latter groups are relatively advantaged.^183,184^ Possible substantive deficiencies are that the model (i) assumes linear effects of maternal age, BMI and height on outcomes, but these effects may be non-linear^20,69,185–188^ (c.f the association between BMI and smoking^94,189^); (ii) ignores the possibility of heterogeneous interactive effects between ethnicity and other causal factors (see below and Tables S7a-l in the Supplementary Information); (iii) does not include known interactions (e.g. risks of low birthweight may increase mainly for older women in deprived areas^185^); (iv) omits many factors that can impact obstetric outcomes, including pregnancy weight gain, immigrant status, rurality, and geopolitical context;^4,25,190,191^ (v) defined its deprivation factor as unidimensional, even though deprivation may have several dimensions;^66,67^ (vi) did not account for multicollinearity resulting from assortative mating. Further studies should test how far these omissions can explain the model’s imperfect fit.

The possibility of heterogeneous interactions between demographic variables and ethnicity presents the greatest threat to the validity of the realistic model. Tables S7a-l in the Supplementary Information provide strong informal evidence of many such interactions in samples of births where parents have the same ethnicity. The most striking of these interactions is that social deprivation has much worse effects on birthweight and the duration of gestation in African Americans than in any other ethnic group (see Table S7b). I have not interpreted these heterogeneous interactive effects, because I had no *a priori* hypotheses about their natures. My use them only to test how far they may account for the imperfect fit of the realistic model. They are so numerous and various that they may account in large part for its imperfect fit. Even so, the fact that the realistic model fitted a 1% sample of the data indicates that that the heterogeneous interactions of ethnicities with other demographic variables are not fatal for the present study. The present findings (Tables S7a-l) should stimulate further studies to confirm and account for the observed heterogeneities.

### Obstetric policy implications

The present study provides a model that can predict the distribution of important outcomes, such as birthweight, for individual pregnancies with any combination of parental risk factors. Such models should yield more accurate prognostic predictions than the simple heuristics in present use^192^ (for example, the current criterion for low birthweight is the same for both sexes – even though male babies are typically 100g heavier^60^). In effect, the present analysis may provide a causal model of the CDC’s observational natality data. There is a recognised need for causal models,^193–195^ and these may guide policies to identify and reduce risks from unnecessary interventions^165,196^ – e.g. possible over-diagnosis of pregnancy hypertension in mothers who pay privately (see above), with its attendant risks of pre-term induction^197^ and consequent low birthweight.

### Wider policy implications

The present findings indicate that ethnicity, deprivation and source of payment for care are important determinants of obstetric outcomes that can have serious, long-term sequelae for both mothers and their babies.^15–18,198^ Obstetrics has relatively well-defined progression and outcomes and so can serve as a model system to disentangle the effects of the above social factors. Extending the present methods may help to elucidate effects of these social factors in other health settings. By analogy with their obstetric effects, the present results imply that policies that reduce deprivation and private insurance and increase state funding of health and social care should help to reduce the overall poor outcomes and high costs of American medicine.^199^

### Data Sharing

All of the data in the present study are freely available from the Centers for Disease Control Natality website.^56,57^ The Supplementary Information includes the full code for data pre-processing and for the statistical analyses and their outputs.

## Supporting information

Addendum 2 - Mplus output for Bayesian estimation of realistic model

Addendum 3a - Mplus output for Baysian estimation of no racism model

Addendum 3b - Mplus output for WLSMV estimation of no racism model

Addendum 4 - Mplus output for Bayesian estimation of realistic model with for 1pct sample

Addendum 1 - R code to read CDC source data and prepare and extract file for Mplus analyses

## Data Availability

All data analysed in this study are available publicly at: https://www.cdc.gov/nchs/data_access/VitalStatsOnline.htm#Births
All data produced in the present study are available upon reasonable request to the author

