## Supplementary Information for "Disentangling effects of ethnicity, deprivation, and payment source on obstetric outcomes in American primigravidae: A structural equation model of observational data"

Table of Contents

| | pages |
|---|---|
| Supplementary Introduction, Methods, Results, Discussion and References | 2 – 18 |
| *Introduction* | 2 |
| *Methods* | 2 – 5 |
| Table S1: correspondence of CDC and analysis variables | 2 |
| Figure S1: details of the Main Report's "realistic" and "no-racism" models | 3 |
| Figure S2: selection of cases for the analyses | 4 |
| *Results* | 5 – 7 |
| Table S2: comparison of selected and excluded births | 5 |
| Table S3: missingness in relation to maternal ethnicity | 5 |
| Table S4: missingness of some variables broken down by maternal ethnicity | 6 |
| Table S5: concordance between maternal and paternal ethnicities | 6 |
| Table S6: direct and total effects in the realistic and no-racism models | 7o |
| Figure S3: distributions of deprivation scores by ethnicity and other factors | 8 |
| Table S7: direct and total effects for each ethnic group, separately | 9 – 18 |
| *Discussion* | 19 – 21 |
| *References* | 22 – 23 |
| Addendum 1: R script to read and preprocess data for Mplus analyses | |
| Addendum 2: Mplus Bayesian output* for the causal model in the Main Report | |
| Input commands | |
| Cross-tabulations of categorical variables | |
| Descriptive statistics of continuous variables | |
| Fit information | |
| Direct effects' path coefficients | |
| Indirect effects' path coefficients | |
| 99% credibility intervals of direct effects | |
| Design matrices, starting values and prior distributions | |
| Addenda 3a-3b: Mplus WLSMV & Bayesian outputs* for the "no racism" model | |
| Input commands | |
| Cross-tabulations of categorical variables | |
| Descriptive statistics of continuous variables | |
| Fit information | |
| Direct effects' path coefficients | |
| Indirect effects' path coefficients | |
| 99% credibility intervals of direct effects | |
| Modification indices | |
| Design matrices and starting values | |
| Addendum 4: Mplus Bayesian analysis of 1% sample | |

* - note that the Bayesian estimations reversed the signs of the loadings for deprivation. Therefore, I re-labelled deprivation from SOCDEP to SOCADV (advantage). Direct path coefficients for SOCADV in these analyses have the opposite sign for the expectation for deprivation.
Mplus commands to perform each analysis appear at the beginning of each Mplus output.

**Introduction**
The main report focuses on effects of ethnicity, deprivation and payment source on obstetric outcomes. Here, I present further details of the sample and analyses and discuss effects of other parental risk factors on each other and on outcomes.

**Methods**
*Variables*
The study used publicly-available natality data from the Centers for Disease Control (CDC).[1] I extracted the study variables from the CDC's file: "Nat2019PublicUS.c20200506.r20200915.txt". The CDC's User Guide ("UserGuide2019-508.pdf")[2] describes these data. Table S1 shows the User Guide's identifiers for the variables, their location in the data file and a brief description of each.

Table S1: correspondence between CDC identifiers and those in the present study

| Variable | CDC name | Position | Brief description | Analysis name |
|---|---|---|---|---|
| Maternal age | MAGER | 75-76 | Mother's single years of age | mage |
| Maternal ethnicity | MRACE31 | 105-106 | Mother's race recode 31 | AFRAMM, OTHM |
| Hispanic mother | MHISPX | 117 | Mother's race/Hispanic origin | MHISP |
| Maternal education | MEDUC | 124 | Mother's education | MEDUC |
| Paternal age | FAGECOMB | 147-148 | Father's combined age | FAGE |
| Paternal ethnicity | FRACE31 | 151-152 | Father's race recode 31 | AFRAMF, OTHF |
| Hispanic father | FHISPX | 162 | Father's race/Hispanic origin | FHISP |
| Paternal education | FEDUC | 163 | Father's education | FEDUC |
| Prior births alive | PRIORLIVE | 171-172 | Prior births now living | *to select primigravidae with singleton pregnancy* |
| Prior births dead | PRIORDEAD | 173-174 | Prior births now dead | |
| Prior terminations | PRIORTERM | 175-176 | Prior other terminations | |
| Plural pregnancy | DPLURAL | 454 | Plurality recode | |
| Beginning of care | PRECARE | 224-225 | Month prenatal care began | PRECAREM |
| Participation in WIC | WIC | 251 | WIC | WIC |
| Pre-pregncy smoking | CIG0 | 253-254 | Cigarettes before pregnancy | *to compute overall smoking before and during pregnancy* |
| 1st trimester smoking | CIG1 | 255-256 | Cigarettes 1st trimester | |
| 2nd trimester smoking | CIG2 | 257-258 | Cigarettes before pregnancy | |
| 3rd trimester smoking | CIG3 | 259-260 | Cigarettes before pregnancy | |
| Mother's height | M_Ht_In | 280-281 | Mother's height in total inches | MHT |
| Mother's BMI | BMI | 283-286 | Body Mass Index | BMI |
| Pre-preg weight | Pwgt_R | 292-294 | Pre-pregnancy weight recode | *To compute BMI* |
| Pre-preg diabetes | RF_PDIAB | 313 | Pre-pregnancy diabetes | predm - *exclusion* |
| Gestational diabetes | RF_GDIAB | 314 | Gestational diabetes | PRGDM |
| Pre-preg hypertensn | RF_PHYPE | 315 | Pre-pregnancy hypertension | prebp - *exclusion* |
| Preg hypertension | RF_GHYPE | 316 | Gestational hypertension | PRGBP |
| Source of payment | PAY_REC | 436 | Payment source for delivery | PAYER |
| Sex of infant | SEX | 475 | Sex of infant | SEXINF |
| Length of gestation | OEGest_Comb | 499-500 | Combined gestation – weeks | OBGEST |
| Birth weight | DBWT | 504-507 | Birthweight – details in grams | BWT |
| Anencephaly | CA_ANEN | 537 | Anencephaly | *exclusion* |

Legend: the CDC's names are from the "User Guide to the 2019 Natality Public Use File".[1] "Position" means the columns that the data occupy in the data file. "Brief Description" is the User Guide's brief description. "Analysis name" is the name in my Mplus analysis (see output). I downloaded some variables in order to exclude them from the model.

Figure 1 in the Main Report shows a schematic diagram of the path model for disentangling effects of ethnicity, deprivation and source of payment. Figure S1 shows the model in detail:

Figure S1: the path diagram for the model in the Main Report

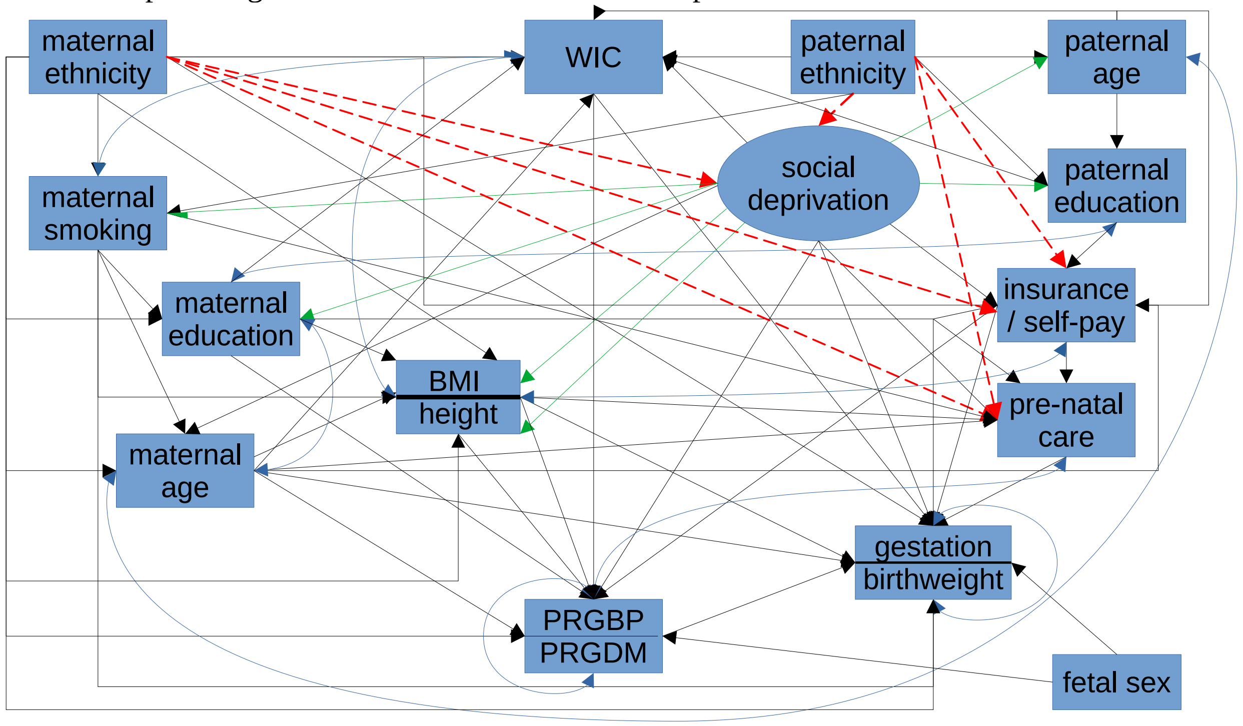

Legend: The path diagram shows the causal paths, factor loadings and non-causal associations between ethnicity, deprivation, payment source (insurance/self-pay) and other variables in the model. Manifest variables are in rectangular boxes, the latent deprivation factor is an oval. Causal paths are straight, single-ended arrows (sometimes with right-angles). Factor loadings are in green; red, dashed arrows are the causal paths that I omitted from the 'no-racism' model; other causal paths that are common to both the 'realistic' and 'no racism' models are black. Non-causal associations are blue, curved double-ended arrows. I combined some factors into horizontally-split boxes, to simplify the diagram. Note that some causal paths partly obscure others – for example, the path from maternal ethnicity to PRGBP overlies the path from maternal age to PRGBP. Figure 2 and Tables 2-5 in the Main Report show the path coefficients for each path on the diagram. Abbreviations: WIC – participation in Supplemental Nutrition Program for Women, Infants and Children; BMI – Body Mass Index; PRGBP – pregnancy hypertension; PRGDM – gestational diabetes mellitus. Note: ethnicity comprises 4 groups – non-Hispanic White (baseline), African American, Hispanic and Other (mostly Asian) ethnicities. The Mplus output in the Supplementary Information may be easier to read and understand than the above path diagram.

*The "no racism" model and separate 'reatlistic' models for each ethnicity*

I assessed the importance of racism by comparing the 'realistic' model with a 'no racism' model that eliminates *direct* effects of parental ethnicity on deprivation and on engagement with obstetric health services (Figure 1 in Main Report and Figure S1, above). Note, however, that the model still allows *in*direct effects of parental ethnicity *via* its (direct) causal effects on parental risk factors. I assessed the adequacy of the realistic model by re-estimating it for each ethnicity separately. In effect, this allows assessment of ethnicity's interactions with other predictors in the realistic model.

*Sample*

I analysed only primigravidae, because prior pregnancy may affect obstetric outcomes.[3–5] I excluded births with incomplete data for any of the study variables. I also excluded: mothers with pre-pregnancy diabetes or hypertension or with extreme values of age (<14 or >45), height (<1.44m or >1.88m), or BMI (<16.5 or >40); babies with extreme values of weight (<1000g or >5000g), or anencephaly, or extreme values of gestational age (<35 weeks or >42 weeks) (see Methods in Main Report and Figure S2). The analyses of each individual ethnicity used only births where both parents were of the same ethnicity.

Figure S2: selection of cases for the analysis

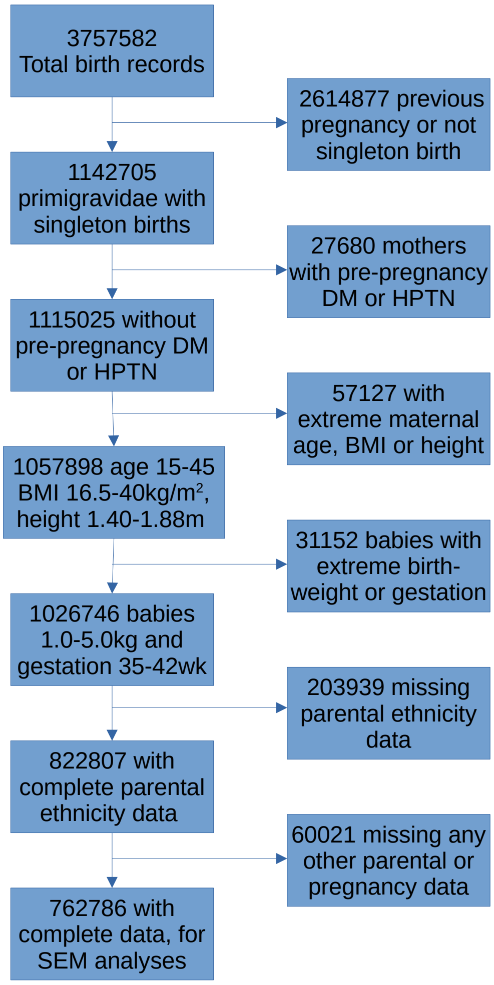

Note: Paternal variables are ethnicity, age and education; maternal variables are ethnicity, age, education, any smoking, participation in WIC, source of payment and gestational month of starting prenatal care. Abbreviations: BMI = body mass index; DM = diabetes mellitus; HPTN = hypertension; wk = weeks; SEM = structural equation modelling.

*Statistical methods*

Simple comparisons between ethnic groups, or between included and excluded cases, used Wilcoxon-Mann-Whitney tests, t-tests, Fisher exact probabilities and $\chi^2$ tests.

*Model estimation and assessment of fit*

Model estimation used Mplus.[6–8] Initial model estimation used the Weighted Least Squares for Mean and Variance (WLSMV) algorithm with ‘theta’ parameterization. I then used the path coefficients from the WLSMV models as starting values for Bayesian re-estimation. These Bayesian analyses used weakly-informative priors centred on zero for most path coefficients, except intercepts, thresholds for categorical variables and residual variances; they used 3 MonteCarlo Markov chains and set the convergence criterion (potential scale reduction) to p=0.005.

I initially assessed the absolute fit of the WLSMV models via their $\chi^2$, Root Mean Square Error of Approximation (RMSEA) and Standardised Root Mean Square Residual (SRMR).[9] However, all these fit indices are imperfect.[10] Therefore, I re-analysed the models using Bayesian estimation of a random 1% sample of the data (N~7800) and assessed this fit using $\chi^2$. I also estimated a “no racism” model (see Figure 1 in Main Report and Supplementary Figure 1) and compared it formally with the ‘realistic’ model using the Satorra-Bentler $\chi^2$ test[6] (see Main Report).

**Results**

*Exclusions*

The excluded cases differed from the study sample for *every* variable in the analysis (Table S2; all ps<$10^{-10}$ via Wilcoxon-Mann-Whitney or Kolmogorov-Smirnov 2-sample tests or Fisher exact probability) including variables which appear identical in Table S2. Note that almost all risk factors and all outcomes were worse in the excluded cases. The most commonly missing variables were paternal ethnicity, education and age (Table S2).

Table S2: the characteristics of the study sample and of excluded cases

| Variable | % missing | Included in study sample | Excluded from study sample |
|---|---|---|---|
| Number of mothers | 0 | 762786 | 263960 |
| Maternal ethnicity | 2.4 | 977636 | 24555 |
| Paternal ethnicity | 19.3 | 828279 | 198467 |
| Smoking mothers (%) | 2.7 | 4.9 | 8.0 |
| Cigs/day (smokers only) | - | 5 (2 – 10) | 5 (2.5 – 10) |
| Maternal Education* | 3.6 | 5 (3 – 6) | 3 (3 – 4) |
| Paternal Education* | 15.4 | 4 ( 3 – 6) | 3 (3 – 5) |
| Maternal age (y) | 2.4 | 27 (23 – 31) | 23 ( 20 – 28) |
| Paternal age (y) | 13.3 | 29 (25 – 33) | 28 (23 – 33) |
| Maternal height (m) | 2.4 | 1.63 (1.57 – 1.68) | 1.63 (1.57 – 1.68) |
| Maternal BMI (kg/m$^2$) | 2.4 | 24.4 (21.6 – 28.5) | 24.8 (21.7 – 29.1.0) |
| WIC participant (%) | 3.4 | 26.0 | 49.2 |
| Privately insured/self-payer (%) | 4.3 | 67.8 | 39.2 |
| Pre-natal care (month) | 6.0 | 2 (2 – 3) | 3 (2 – 5) |
| Pregnancy high BP (%) | 2.4 | 8.5 | 8.8 |
| Gestational diabetes (%) | 2.4 | 4.3 | 5.4 |
| Gestation (weeks) | 2.4 | 39 (38 – 40) | 39 (38 – 40) |
| Birthweight (kg) | 2.4 | 3.31 (3.01 – 3.60) | 3.24 (2.94 – 3.54) |

Legend: the values are counts (numbers of parents), percentages, or medians – with inter-quartile ranges in brackets).

Missing data (apart from ethnicity) were more common for AA and Other-ethnicity mothers:

Table S3: Missingness of any other variable, in relation to non-missing maternal ethnicity

| Other data | Maternal ethnicity | nHEA | African American | Hispanic | Other |
|---|---|---|---|---|---|
| Not missing | | 451267 | 89356 | 136847 | 85316 |
| Missing | | 112091 | 57475 | 77535 | 16859 |
| Odds ratio | | 1 (Reference) | **2.59** | **2.28** | **0.80** |

Variables apart from maternal ethnicity showed different patterns of missingness in relation to non-missing maternal ethnicity. Compared with nHEA mothers, paternal variables were missing more frequently for African American and Hispanic mothers (Table S4), but less frequently for Other-ethnicity mothers. In contrast, missingness of maternal variables, participation in WIC and source of payment was *apparently* less prevalent in mothers of all ethnic-minorities (Table S4). This is probably because electronic birth records that had missing maternal ethnicity also had missing data for paternal variables, so that missingness for maternal variables was only a small proportion of total missingness for those cases.

Table S4: numbers of missing values of different variables, broken down by maternal ethnicity

| Variable | Missing /OR | nHEA | AA | Hispanic | Other | $\chi^2$ (3df) |
|---|---|---|---|---|---|---|
| Paternal age | No | 499600 | 107679 | 214382 | 97428 | 20692 |
| | Yes | 63758 | 39152 | 28723 | 4747 | |
| | OR | Ref. | **2.85** | **1.21** | **0.38** | |
| Paternal education | No | 489909 | 103909 | 181197 | 94059 | 28402 |
| | Yes | 73449 | 42922 | 33185 | 8116 | |
| | OR | Ref. | **2.75** | **1.22** | **0.58** | |
| Maternal education | No | 533012 | 145332 | 211891 | 99150 | 11663 |
| | Yes | 30346 | 1499 | 2491 | 3025 | |
| | OR | Ref. | **0.18** | **0.21** | **0.54** | |
| Maternal smoking | No | 536893 | 146039 | 213852 | 101793 | 17893 |
| | Yes | 26465 | 792 | 530 | 382 | |
| | OR | Ref. | **0.11** | **0.05** | **0.08** | |
| Participation in WIC | No | 533892 | 145231 | 212675 | 100418 | 13575 |
| | Yes | 29466 | 1600 | 1707 | 1757 | |
| | OR | Ref. | **0.20** | **0.15** | **0.32** | |
| Source of payment | No | 529024 | 143752 | 209510 | 100160 | 9596 |
| | Yes | 34334 | 3079 | 4872 | 2015 | |
| | OR | Ref. | **0.33** | **0.36** | **0.31** | |

Legend: The table shows three rows for each variable, broken down by maternal ethnicity. For each variable, the first row shows the numbers of cases with no missing values; the second row shows the number of cases with missing values and the third row shows the odds ratio for comparing each minority ethnicity with non-Hispanic European American (nHEA) mothers. Abbreviations: AA = African American; BMI = body mass index; WIC = Supplemental Food Program for Women Infants and Children; OR = odds ratio; Ref. = reference. Bold values differ significantly from nHEA

The above comparisons (Tables S2 – S3) indicate clearly that the missing data are “not missing at random” (NMAR). In fact, maternal ethnicity is a strong predictor of missingness. Note, however, that these results are uncertain, because they depend on missingness of both maternal ethnicity and the other variables, which may be inter-dependent. The reasons for this missingness are unknown.

Recorded maternal and paternal ethnicities tend to be concordant (Cohen’s kappa = 0.72, Table S5).

Table S5: Concordance between maternal and paternal ethnicities

| Maternal ethnicity | Paternal ethnicity: | nHEA | African American | Hispanic | Other |
|---|---|---|---|---|---|
| nHEA | | 500423 | 19092 | 31432 | 12411 |
| African American | | 50840 | 87292 | 7678 | 1021 |
| Hispanic | | 61547 | 8060 | 140839 | 3936 |
| Other | | 26631 | 2584 | 4847 | 68113 |

The table coarsens values for AA and Other ethnicities, and recodes parents reporting only two ‘races’ (CDC ‘mrace’ codes 6-10, 13 and 15) as AA or Other.

This concordance means there is some collinearity in parental ethnicities. Potentially, parental ethnicities could interact when causing other risk factors or outcomes. The present analyses do not attempt to define or quantify such interactions.

**The “no racism” model**

The 'no-racism' model did not fit the data well (see Main Report). Therefore, I only contrast it with the ‘realistic’ model, and with the raw data. The realistic and no-racism models showed different direct effects of parental ethnicities on obstetric risk factors – especially age and education (see Table S6a-S6b and Mplus outputs). Specifically, *direct* effects of ethnicity tended to be more detrimental for AA and Hispanic parents, but *total* effects of ethnicity were generally similar in the two models (Table S6c and Mplus outputs).

Table S6a: Direct and total effects of parental ethnicity on parental age and education in the realistic and no-racism models

| Variable | Maternal age | | | | Paternal age | | | | Maternal Education | | | | Paternal Education | | | |
|---|---|---|---|---|---|---|---|---|---|---|---|---|---|---|---|---|
| Effect type | Direct | | Total | | Direct | | Total | | Direct | | Total | | Direct | | Total | |
| model | realistic | noracism | realistic | noracism | realistic | noracism | realistic | noracism | realistic | noracism | realistic | noracism | realistic | noracism | realistic | noracism |
| AFAMM | -0.1 | -3.3 | -4.5 | -4.4 | - | - | -3.4 | - | 0.6 | -2.1 | -3.4 | -3.5 | - | - | -4.7 | - |
| MHISP | 1.2 | -1.9 | -8.5 | -3.8 | - | - | -7.6 | - | -4.0 | -6.6 | -13.9 | -9 | - | - | -10.4 | - |
| OTHM | -0.2 | 9.5 | 9.8 | 5.5 | - | - | 8.2 | - | -3.9 | 10.4 | 8.8 | 5.4 | - | - | 11.2 | - |
| AFAMF | - | - | -14 | -2.7 | 1.5 | -3.6 | -9.8 | -3.6 | - | - | -16.4 | -3.3 | 2.2 | - | -13.2 | -1.4 |
| FHISP | - | - | -13.5 | -4.4 | 0.3 | -6.1 | -10.4 | -6.1 | - | - | -15.1 | -5.4 | -4.2 | - | -18.8 | -3 |
| OTHF | - | - | 0.3 | 0.9 | 2.7 | 7.3 | 3.0 | 7.3 | - | - | 0.7 | 1.1 | 2.6 | - | 2.9 | 2.2 |

Table S6b: Direct and total effects of parental ethnicity on maternal smoking, BMI, source of payment and WIC in the realistic and no-racism models

| Variable | Maternal smoking | | | | Pre-pregnancy BMI | | | | Source of payment | | | | Participation in WIC | | | |
|---|---|---|---|---|---|---|---|---|---|---|---|---|---|---|---|---|
| Effect type | Direct | | Total | | Direct | | Total | | Direct | | Total | | Direct | | Total | |
| model | realistic | noracism | realistic | noracism | realistic | noracism | realistic | noracism | realistic | noracism | realistic | noracism | realistic | noracism | realistic | noracism |
| AFAMM | -17.0 | -6.6 | -13.9 | -6.6 | 0.8 | 4.9 | 2.6 | 3.4 | -6.8 | - | -11.3 | - | 7.1 | 9.3 | 11.5 | 7.6 |
| MHISP | -22.8 | -11.0 | -15.8 | -11.0 | -0.6 | 4.9 | 1.9 | 1.3 | -4.6 | - | -14.6 | - | 5.4 | 11.9 | 14.9 | 7.7 |
| OTHM | -8.5 | -22.6 | -16.1 | -22.6 | -8.3 | -25.1 | -11.8 | -10.8 | -9.2 | - | 1.7 | - | 7.0 | -3.5 | -3.5 | -1.0 |
| AFAMF | -6.1 | -15 | 4.3 | -15.0 | - | - | 4.0 | 2.5 | -3.2 | - | -18.3 | -1.4 | 4.7 | 9.4 | 19.0 | 7.9 |
| FHISP | -15.2 | -24.8 | -5.3 | -24.8 | - | - | 4.1 | 4.1 | -2.7 | - | -16.6 | -3.0 | 4.8 | 12.5 | 18.5 | 10.0 |
| OTHF | -6.0 | 5.1 | -6.3 | 5.1 | - | - | 0.1 | -0.9 | -3.1 | - | -3.0 | 2.2 | 4.7 | 1.7 | 4.4 | 2.3 |

Table S6c: Direct and total effects of parental ethnicity on pregnancy and birth outcomes in the realistic and no-racism models

| Variable | Pregnancy hypertension | | | | Gestational diabetes | | | | Duration of gestation | | | | Birth weight | | | |
|---|---|---|---|---|---|---|---|---|---|---|---|---|---|---|---|---|
| Effect type | Direct | | Total | | Direct | | Total | | Direct | | Total | | Direct | | Total | |
| model | realistic | noracism | realistic | noracism | realistic | noracism | realistic | noracism | realistic | noracism | realistic | noracism | realistic | noracism | realistic | noracism |
| AFAMM | 1.7 | 7.4 | 1.7 | 1.6 | -0.3 | -0.7 | -1.1 | -0.9 | -2.1 | -2.8 | -2.2 | -2.3 | -9.8 | -8.8 | -8.9 | -8.9 |
| MHISP | -4.7 | 3.0 | -4.8 | -5.5 | 0.9 | 0.5 | 0.9 | 1.2 | -1.4 | -2.3 | -0.9 | -0.6 | -0.4 | 0.7 | -2.0 | -1.8 |
| OTHM | -1.6 | -22.8 | -5.9 | -5.1 | 11.5 | 11.8 | 11.0 | 10.6 | -3.2 | -0.6 | -3.3 | -3.7 | -2.6 | -5.1 | -5.7 | -5.8 |
| AFAMF | -3.0 | -0.4 | -1.7 | -1.9 | -3.5 | -2.7 | -4.1 | -3.5 | -2.7 | -3.4 | -2.1 | -1.7 | -4.4 | -5.5 | -4.5 | -3.5 |
| FHISP | -2.6 | -0.6 | -1.3 | -1.1 | -1.2 | -0.2 | -2.3 | -1.5 | -1.6 | -2.4 | -1.4 | -1.1 | -4.2 | -5.7 | -3.6 | -2.6 |
| OTHF | -4.3 | -2.5 | -4.8 | -5.4 | 3.0 | 2.2 | 2.8 | 2.7 | -0.7 | -0.7 | 0.6 | 0.8 | -7.9 | -6.7 | -6.5 | -6.5 |
| SOCDEP | 17.3 | 17.4 | 8.5 | 14.1 | 4.1 | 1.1 | 1.4 | 0.5 | -9.0 | -20.2 | -6.2 | -7.2 | -6.9 | 16.2 | -6.2 | -5.3 |

Legend: the values in tables 6a-6c are doubly-standardised path coefficients from the realistic and no-racism models for direct and total effects of parental ethnicities and social deprivation on risk factors, financial indicators and obstetric outcomes. Values in green or red show where the no-racism model predicts better or worse outcomes than the realistic model.

---

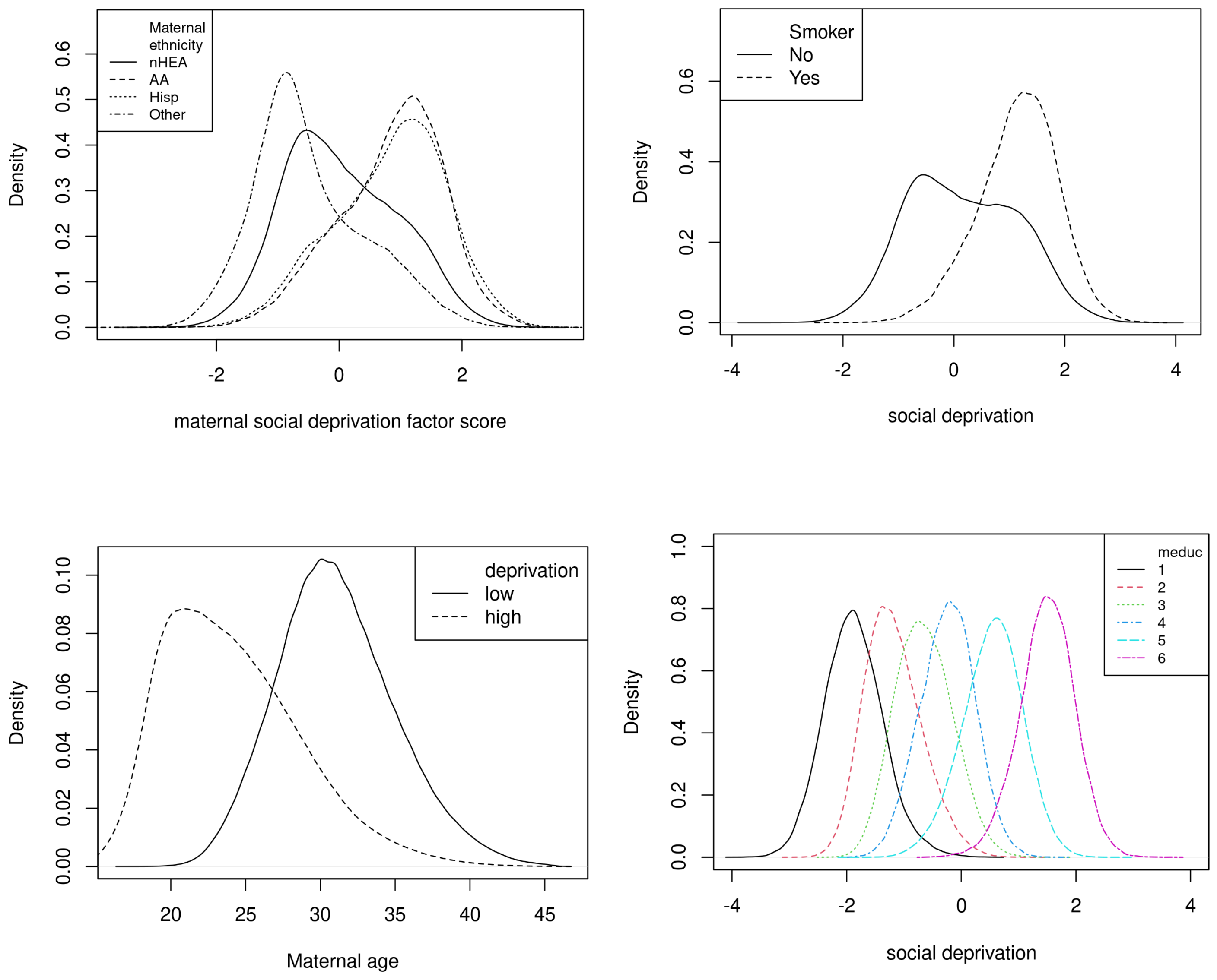

Figure S3 (a-d): Associations between factor scores of social deprivation and maternal demographic characteristics.

(a) (top left): distributions of deprivation factor scores in 4 ethnic groups – non-Hispanic European American (nHEA), African American (AA), Hispanic (Hisp) and Other (mostly Asian).

(b) (top right) distributions of deprivation factor scores in non-smokers and smokers

(c) (bottom left) distributions of maternal age in mothers whose deprivation scores were low (<0) or high (>0).

(d) (bottom right) distributions of deprivation scores in relation to maternal educational level (recoded from CDC categorisation: 1 = none through 12th grade; 2 = high school graduate; 3 = some college credit, but not a degree; 4 = Associate degree; 5 = Bachelor's degree or Master's degree; 6 = Doctorate or Professional degree.

Table S7a: Loadings of parental demographic variables on the social deprivation factor, separately for each ethnic group

| Ethnicity: | Non-Hispanic European American | | | African-American | | | Hispanic | | | Other (mostly Asian) | | |
|---|---|---|---|---|---|---|---|---|---|---|---|---|
| loading | Intercept | low 95%CI | high 95%CI | Intercept | low 95%CI | high 95%CI | Intercept | low 95%CI | high 95%CI | Intercept | low 95%CI | high 95%CI |
| FAGE | -0.645 | -0.650 | -0.640 | -0.773 | -0.787 | -0.759 | -0.712 | -0.724 | -0.700 | -0.333 | -0.345 | -0.321 |
| FEDUC | -1.213 | -1.241 | -1.186 | -0.822 | -0.860 | -0.790 | -0.987 | -1.030 | -0.943 | -1.097 | -1.183 | -1.027 |
| MEDUC | -1.407 | -1.438 | -1.377 | -1.258 | -1.304 | -1.214 | -1.003 | -1.029 | -0.977 | -1.198 | -1.261 | -1.136 |
| MHT | -0.117 | -0.120 | -0.115 | -0.088 | -0.094 | -0.082 | -0.133 | -0.137 | -0.128 | -0.092 | -0.098 | -0.087 |
| CIG0 | 0.645 | 0.633 | 0.657 | 0.204 | 0.170 | 0.240 | 0.071 | 0.036 | 0.106 | 0.607 | 0.540 | 0.675 |
| BMI | 0.577 | 0.543 | 0.611 | 0.607 | 0.494 | 0.730 | 0.287 | 0.254 | 0.320 | 0.258 | 0.197 | 0.327 |

Legend: Values are mean loadings from Bayesian estimation, together with their lower and upper 95% credibility intervals (CI). Key: FAGE = paternal age; FEDUC = paternal education; MEDUC = maternal education; MHT = maternal height; CIG0 = any previous maternal smoking; BMI = pre-pregnancy maternal BMI. For ethnic minorities, loadings in red are stronger than those in the non-Hispanic European American (nHEA) group (their 95% CIs do not overlap); loadings in green are weaker.

Table S7b: effects of social deprivation on maternal age, indicators of obstetric care and obstetric outcomes, separately for each ethnic group

| Ethnicity | Non-Hispanic European American | | | African-American | | | Hispanic | | | Other (mostly Asian) | | |
|---|---|---|---|---|---|---|---|---|---|---|---|---|
| SOCDEP → | Intercept | low 95%CI | high 95%CI | Intercept | low 95%CI | high 95%CI | Intercept | low 95%CI | high 95%CI | Intercept | low 95%CI | high 95%CI |
| MAGE | -0.699 | -0.705 | -0.693 | -0.788 | -0.800 | -0.775 | -0.704 | -0.714 | -0.695 | -0.440 | -0.458 | -0.422 |
| PAYER | -1.209 | -1.260 | -1.168 | -0.941 | -1.000 | -0.886 | -1.112 | -1.209 | -1.023 | -1.429 | -1.674 | -1.208 |
| WIC | 0.970 | 0.911 | 1.033 | 1.335 | 1.036 | 1.653 | 0.635 | 0.573 | 0.701 | 1.376 | 1.040 | 1.808 |
| PRECAREM | 0.030 | 0.005 | 0.054 | -0.511 | -0.769 | -0.300 | 0.097 | 0.043 | 0.154 | -0.240 | -0.442 | -0.072 |
| PRGBP | 0.165 | 0.123 | 0.206 | 0.218 | -0.015 | 0.475 | 0.086 | 0.011 | 0.161 | 0.244 | -0.03 | 0.586 |
| PRGDM | 0.023 | -0.027 | 0.071 | -0.216 | -0.563 | 0.089 | 0.166 | 0.079 | 0.256 | -0.072 | -0.292 | 0.131 |
| OBGEST | -0.077 | -0.108 | -0.044 | **-0.700** | **-1.074** | **-0.438** | 0.018 | -0.034 | 0.070 | -0.153 | -0.361 | 0.015 |
| BWT | -0.018 | -0.029 | -0.008 | **-0.315** | **-0.464** | **-0.205** | -0.018 | -0.036 | 0.000 | -0.027 | -0.089 | 0.029 |

Legend: Values are mean path coefficients from Bayesian estimation, together with their lower and upper 95% credibility intervals (CI). Key: MAGE = maternal age; PAYER = insured or private payer; WIC = participation in WIC; PRECAREM = month of gestation when pre-natal care began; PRGBP = pregnancy hypertension; PRGDM = gestational diabetes; OBGEST = duration of gestation; BWT = birthweight. For ethnic minorities, loadings in red are more adverse than those in the non-Hispanic European American (nHEA) group (their 95% CIs do not overlap); loadings in green are more beneficial. Note that deprivation causes much worse birth outcomes for AA parents than in nHEA parents (highlighted in bold), but deprivation's effects on birth outcomes in Hispanic and Other parents are similar to those for nHEA parents..

Table S7a: Variances of continuous variables for each of ethnic groupings

| Ethnicity: | Non-Hispanic European American | | | African-American | | | Hispanic | | | Other (mostly Asian) | | |
|---|---|---|---|---|---|---|---|---|---|---|---|---|
| Variable | Intercept | low 95%CI | high 95%CI | Intercept | low 95%CI | high 95%CI | Intercept | low 95%CI | high 95%CI | Intercept | low 95%CI | high 95%CI |
| FAGE | 1.028 | 1.021 | 1.035 | 1.425 | 1.404 | 1.447 | 1.19 | 1.173 | 1.207 | 1.071 | 1.058 | 1.085 |
| MAGE | 0.591 | 0.586 | 0.596 | 0.598 | 0.583 | 0.614 | 0.666 | 0.654 | 0.677 | 0.641 | 0.631 | 0.651 |
| MHT | 0.443 | 0.441 | 0.445 | 0.461 | 0.456 | 0.466 | 0.379 | 0.376 | 0.383 | 0.339 | 0.335 | 0.343 |
| BMI | 0.907 | 0.898 | 0.916 | 1.049 | 1.007 | 1.087 | 0.969 | 0.958 | 0.980 | 0.623 | 0.609 | 0.635 |
| PRECAREM | 1.275 | 1.27 | 1.281 | 2.206 | 2.161 | 2.244 | 2.278 | 2.257 | 2.299 | 1.949 | 1.921 | 1.975 |
| OBGEST | 1.423 | 1.416 | 1.431 | 1.438 | 1.368 | 1.483 | 1.425 | 1.409 | 1.440 | 1.381 | 1.359 | 1.402 |
| BWT | 0.186 | 0.185 | 0.187 | 0.180 | 0.169 | 0.188 | 0.181 | 0.179 | 0.183 | 0.173 | 0.171 | 0.175 |

Legend: Intercepts are variances from Bayesian estimation, together with their lower and upper 95% credibility intervals. Variances that are greater in ethnic minorities (95%CIs do not overlap those of nHEA) are in red; those that are smaller are in green. Key: FAGE = paternal age; MAGE = maternal age; MHT = maternal height; BMI = pre-pregnancy maternal BMI; PRECAREM = month of gestation when pre-natal care began; OBGEST = duration of gestation (obstetric estimate); BWT = birthweight.

Table S7b: Miscellaneous effects of different demographic variables on each other

| Ethnicity | | Non-Hispanic European American | | | African-American | | | Hispanic | | | Other (mostly Asian) | | |
|---|---|---|---|---|---|---|---|---|---|---|---|---|---|
| Cause | effect | Intercept | low 95%CI | high 95%CI | Intercept | low 95%CI | high 95%CI | Intercept | low 95%CI | high 95%CI | Intercept | low 95%CI | high 95%CI |
| FAGE | FEDUC | 0.003 | -0.006 | 0.011 | 0.071 | 0.059 | 0.082 | -0.063 | -0.079 | -0.048 | 0.012 | -0.002 | 0.025 |
| CIG0 | MAGE | 0.011 | 0.006 | 0.017 | 0.019 | 0.002 | 0.036 | -0.027 | -0.047 | -0.007 | 0.021 | -0.004 | 0.046 |
| CIG0 | MEDUC | -0.120 | -0.134 | -0.107 | -0.113 | -0.146 | -0.079 | -0.069 | -0.099 | -0.039 | -0.001 | -0.055 | 0.053 |
| MEDUC | BMI | 0.126 | 0.114 | 0.138 | 0.226 | 0.187 | 0.268 | 0.084 | 0.070 | 0.098 | 0.088 | 0.064 | 0.114 |
| MAGE | BMI | 0.216 | 0.207 | 0.226 | 0.354 | 0.311 | 0.398 | 0.199 | 0.185 | 0.213 | 0.047 | 0.032 | 0.063 |
| CIG0 | BMI | -0.035 | -0.043 | -0.026 | 0.040 | 0.014 | 0.067 | 0.023 | -0.003 | 0.050 | 0.059 | 0.027 | 0.090 |

Legend: Intercepts are path coefficients from causal factors to effects, from Bayesian estimation, together with their lower and upper 95% credibility intervals. Coefficients that are higher in ethnic minorities (95%CIs do not overlap those of nHEA) are in red; those that are lower are in green. Key: as in Table S7a-b..

Table S7c: effects of paternal and maternal age and education on source of payment and participation in WIC

| Ethnicity | Non-Hispanic European American | | | African-American | | | Hispanic | | | Other (mostly Asian) | | |
|---|---|---|---|---|---|---|---|---|---|---|---|---|
| path | Intercept | lo 95%CI | hi 95%CI | Intercept | lo 95%CI | hi 95%CI | Intercept | lo 95%CI | hi 95%CI | Intercept | lo 95%CI | hi 95%CI |
| FAGE→ PAYER | -0.010 | -0.020 | 0.000 | -0.016 | -0.030 | -0.003 | -0.030 | -0.052 | -0.009 | 0.056 | 0.036 | 0.074 |
| FEDUC → PAYER | -0.092 | -0.112 | -0.074 | -0.002 | -0.030 | 0.023 | -0.094 | -0.132 | -0.058 | 0.010 | -0.082 | 0.092 |
| MAGE → WIC | 0.008 | -0.013 | 0.028 | 0.169 | 0.085 | 0.259 | -0.039 | -0.061 | -0.016 | 0.071 | -0.027 | 0.199 |
| MEDUC → WIC | -0.146 | -0.160 | -0.131 | 0.123 | 0.039 | 0.214 | -0.064 | -0.086 | -0.040 | -0.035 | -0.087 | 0.024 |

Legend: Intercepts are path coefficients from causal factors to effects, from Bayesian estimation, together with their lower and upper 95% credibility intervals. Coefficients that are higher in ethnic minorities (compared with nHEA) are in red; those that are lower are in green. Key: as in Tables S7a-b.

Table S7d: effects of demographic factors, payment source and participation in WIC on initiation of pre-natal care (PNC)

| Ethnicity | Non-Hispanic European American | | | African-American | | | Hispanic | | | Other (mostly Asian) | | |
|---|---|---|---|---|---|---|---|---|---|---|---|---|
| x → PNC | Intercept | lo 95%CI | hi 95%CI | Intercept | lo 95%CI | hi 95%CI | Intercept | lo 95%CI | hi 95%CI | Intercept | lo 95%CI | hi 95%CI |
| MAGE | -0.019 | -0.026 | -0.013 | -0.170 | -0.239 | -0.111 | -0.050 | -0.068 | -0.031 | -0.096 | -0.116 | -0.078 |
| MEDUC | -0.034 | -0.041 | -0.027 | -0.189 | -0.254 | -0.135 | -0.061 | -0.079 | -0.043 | -0.09 | -0.130 | -0.054 |
| PAYER | -0.102 | -0.110 | -0.094 | -0.183 | -0.210 | -0.155 | -0.068 | -0.090 | -0.045 | -0.169 | -0.206 | -0.133 |
| CIG0 | 0.023 | 0.016 | 0.031 | -0.032 | -0.068 | 0.004 | 0.028 | -0.009 | 0.066 | 0.034 | -0.024 | 0.093 |
| BMI | -0.027 | -0.032 | -0.022 | 0.059 | 0.021 | 0.108 | -0.051 | -0.063 | -0.040 | 0.061 | 0.032 | 0.096 |
| WIC | -0.036 | -0.045 | -0.028 | 0.034 | -0.026 | 0.106 | -0.110 | -0.128 | -0.092 | -0.012 | -0.060 | 0.050 |

Legend: Intercepts are path coefficients for causes of pre-natal care (PNC), from Bayesian estimation, together with their lower and upper 95% credibility intervals. Coefficients that delay beginning PNC in ethnic minorities (95%CIs do not overlap those of nHEA) are in red; those that are cause earlier initiation of PNC are in green. Key: as in Tables S7a-b.

Table S7e: effects of demographic and care variables and fetal sex on pregnancy hypertension, separately in each ethnic group.

| Ethnicity: | Non-Hispanic European American | | | African-American | | | Hispanic | | | Other (mostly Asian) | | |
|---|---|---|---|---|---|---|---|---|---|---|---|---|
| x → PRGBP | Intercept | lo 95%CI | hi 95%CI | Intercept | lo 95%CI | hi 95%CI | Intercept | lo 95%CI | hi 95%CI | Intercept | lo 95%CI | hi 95%CI |
| CIG0 | 0.017 | 0.005 | 0.029 | 0.010 | -0.029 | 0.049 | -0.026 | -0.077 | 0.024 | 0.008 | -0.084 | 0.096 |
| FAGE | -0.020 | -0.028 | -0.013 | -0.006 | -0.021 | 0.009 | -0.002 | -0.018 | 0.013 | -0.044 | -0.067 | -0.021 |
| MAGE | 0.031 | 0.019 | 0.042 | 0.072 | 0.009 | 0.141 | 0.029 | 0.005 | 0.053 | 0.121 | 0.086 | 0.158 |
| MEDUC | 0.032 | 0.021 | 0.044 | 0.038 | -0.024 | 0.104 | 0.005 | -0.019 | 0.027 | 0.020 | -0.037 | 0.090 |
| MHT | 0.037 | 0.029 | 0.045 | 0.017 | -0.003 | 0.038 | 0.013 | -0.008 | 0.034 | -0.022 | -0.055 | 0.011 |
| BMI | 0.211 | 0.203 | 0.218 | 0.121 | 0.080 | 0.158 | 0.158 | 0.144 | 0.172 | 0.235 | 0.184 | 0.278 |
| PAYER | 0.044 | 0.030 | 0.058 | 0.020 | -0.013 | 0.052 | -0.004 | -0.032 | 0.026 | 0.000 | -0.061 | 0.063 |
| WIC | -0.036 | -0.051 | -0.022 | -0.022 | -0.084 | 0.031 | -0.004 | -0.026 | 0.018 | -0.062 | -0.148 | 0.004 |
| SEXINF | 0.025 | 0.014 | 0.036 | 0.025 | -0.001 | 0.052 | 0.037 | 0.013 | 0.062 | 0.008 | -0.029 | 0.045 |

Legend: Intercepts are path coefficients from causal factors to pregnancy hypertension (PRGBP), from Bayesian estimation, together with their lower and upper 95% credibility intervals. Coefficients that show more PRGBP in ethnic minorities (95%CIs do not overlap those of nHEA) are in red; those that show less PRGBP are in green. Key: as in Tables S7a-b.

Table S7f: effects of demographic and care variables and fetal sex on gestational diabetes, separately in each ethnic group.

| Ethnicity: | Non-Hispanic European American | | | African-American | | | Hispanic | | | Other (mostly Asian) | | |
|---|---|---|---|---|---|---|---|---|---|---|---|---|
| x → PRGDM | Intercept | lo 95%CI | hi 95%CI | Intercept | lo 95%CI | hi 95%CI | Intercept | lo 95%CI | hi 95%CI | Intercept | lo 95%CI | hi 95%CI |
| CIG0 | 0.054 | 0.040 | 0.069 | 0.033 | -0.021 | 0.086 | 0.051 | -0.005 | 0.106 | -0.002 | -0.078 | 0.074 |
| FAGE | 0.011 | 0.002 | 0.020 | 0.024 | 0.004 | 0.043 | 0.005 | -0.013 | 0.022 | 0.011 | -0.006 | 0.028 |
| MAGE | 0.155 | 0.142 | 0.169 | 0.114 | 0.024 | 0.193 | 0.235 | 0.208 | 0.263 | 0.173 | 0.147 | 0.198 |
| MEDUC | -0.008 | -0.022 | 0.005 | -0.066 | -0.155 | 0.016 | 0.025 | -0.002 | 0.052 | -0.018 | -0.062 | 0.024 |
| MHT | -0.124 | -0.134 | -0.113 | -0.046 | -0.075 | -0.017 | -0.044 | -0.068 | -0.020 | -0.158 | -0.183 | -0.134 |
| BMI | 0.201 | 0.191 | 0.210 | 0.186 | 0.136 | 0.246 | 0.194 | 0.177 | 0.210 | 0.241 | 0.208 | 0.276 |
| PAYER | 0.026 | 0.009 | 0.043 | -0.040 | -0.086 | 0.005 | 0.036 | 0.002 | 0.070 | -0.051 | -0.098 | -0.006 |
| WIC | 0.021 | 0.002 | 0.039 | 0.060 | -0.011 | 0.141 | 0.000 | -0.027 | 0.027 | 0.017 | -0.030 | 0.067 |
| SEXINF | 0.020 | 0.007 | 0.034 | 0.032 | -0.006 | 0.068 | 0.002 | -0.026 | 0.031 | 0.019 | -0.007 | 0.046 |

Legend: Intercepts are path coefficients from causal factors to gestational diabetes (PRGDM), from Bayesian estimation, together with their lower and upper 95% credibility intervals. Coefficients that show more PRGDM in ethnic minorities (95%CIs do not overlap those of nHEA) are in red; those that show less PRGDM are in green. Key: as in Tables S7a-b.

Table S7g: effects of demographic and care variables and fetal sex on birthweight, separately in each ethnic group.

| Ethnicity: | Non-Hispanic European American | | | African-American | | | Hispanic | | | Other (mostly Asian) | | |
|---|---|---|---|---|---|---|---|---|---|---|---|---|
| x → BWT | Intercept | lo 95%CI | hi 95%CI | Intercept | lo 95%CI | hi 95%CI | Intercept | lo 95%CI | hi 95%CI | Intercept | lo 95%CI | hi 95%CI |
| CIG0 | -0.030 | -0.034 | -0.027 | -0.050 | -0.064 | -0.036 | -0.013 | -0.025 | -0.002 | 0.019 | 0.000 | 0.037 |
| FAGE | -0.009 | -0.011 | -0.008 | 0.000 | -0.004 | 0.005 | 0.001 | -0.003 | 0.005 | -0.005 | -0.010 | -0.001 |
| MAGE | -0.019 | -0.022 | -0.016 | -0.084 | -0.123 | -0.054 | -0.006 | -0.012 | 0.000 | 0.004 | -0.004 | 0.011 |
| MEDUC | -0.005 | -0.008 | -0.002 | -0.073 | -0.110 | -0.045 | -0.005 | -0.010 | 0.001 | -0.010 | -0.022 | 0.002 |
| MHT | 0.135 | 0.133 | 0.137 | 0.097 | 0.091 | 0.103 | 0.119 | 0.114 | 0.124 | 0.138 | 0.131 | 0.144 |
| BMI | 0.072 | 0.070 | 0.074 | 0.095 | 0.074 | 0.125 | 0.068 | 0.065 | 0.072 | 0.077 | 0.068 | 0.087 |
| PAYER | 0.000 | -0.003 | 0.004 | -0.030 | -0.041 | -0.020 | 0.000 | -0.007 | 0.007 | -0.006 | -0.018 | 0.007 |
| WIC | -0.008 | -0.012 | -0.004 | 0.065 | 0.034 | 0.106 | 0.010 | 0.005 | 0.015 | 0.010 | -0.002 | 0.025 |
| PNC | -0.002 | -0.003 | 0.000 | -0.008 | -0.016 | -0.001 | -0.002 | -0.004 | 0.000 | 0.002 | -0.001 | 0.005 |
| PRGBP | -0.085 | -0.088 | -0.083 | -0.078 | -0.092 | -0.061 | -0.084 | -0.089 | -0.079 | -0.068 | -0.076 | -0.060 |
| PRGDM | -0.009 | -0.012 | -0.006 | 0.024 | 0.002 | 0.041 | 0.004 | -0.002 | 0.01 | -0.025 | -0.031 | -0.019 |
| SEXINF | 0.124 | 0.121 | 0.127 | 0.118 | 0.111 | 0.125 | 0.104 | 0.099 | 0.110 | 0.088 | 0.081 | 0.096 |

Legend: Intercepts are path coefficients from causal factors to birthweight (BWT), from Bayesian estimation, together with their lower and upper 95% credibility intervals. Coefficients for variables that increase BWT in ethnic minorities (95%CIs do not overlap those of nHEA) are in green; those that decrease BWT are in red. Key: as in Tables S7a-b.

Table S7h: effects of demographic and care variables and fetal sex on duration of gestation, separately in each ethnic group.

| Ethnicity: | Non-Hispanic European American | | | African-American | | | Hispanic | | | Other (mostly Asian) | | |
|---|---|---|---|---|---|---|---|---|---|---|---|---|
| x → GEST | Intercept | lo 95%CI | hi 95%CI | Intercept | lo 95%CI | hi 95%CI | Intercept | lo 95%CI | hi 95%CI | Intercept | lo 95%CI | hi 95%CI |
| CIG0 | -0.005 | -0.014 | 0.005 | -0.047 | -0.084 | -0.011 | 0.006 | -0.028 | 0.041 | 0.045 | -0.013 | 0.106 |
| FAGE | -0.006 | -0.012 | 0.000 | 0.018 | 0.005 | 0.031 | 0.005 | -0.006 | 0.015 | -0.008 | -0.021 | 0.006 |
| MAGE | 0.012 | 0.003 | 0.021 | -0.185 | -0.279 | -0.111 | 0.035 | 0.017 | 0.053 | -0.009 | -0.031 | 0.013 |
| MEDUC | -0.013 | -0.022 | -0.004 | -0.163 | -0.259 | -0.096 | 0.013 | -0.003 | 0.029 | -0.014 | -0.055 | 0.021 |
| MHT | 0.061 | 0.054 | 0.068 | 0.028 | 0.011 | 0.046 | 0.075 | 0.061 | 0.090 | 0.070 | 0.050 | 0.089 |
| BMI | 0.090 | 0.084 | 0.096 | 0.202 | 0.152 | 0.275 | 0.088 | 0.077 | 0.098 | 0.108 | 0.081 | 0.142 |
| PAYER | -0.015 | -0.025 | -0.004 | -0.075 | -0.105 | -0.045 | 0.006 | -0.014 | 0.026 | -0.017 | -0.054 | 0.021 |
| WIC | -0.021 | -0.033 | -0.010 | 0.153 | 0.084 | 0.253 | 0.034 | 0.018 | 0.050 | 0.046 | 0.006 | 0.097 |
| PNC | 0.053 | 0.049 | 0.057 | 0.033 | 0.013 | 0.049 | 0.038 | 0.032 | 0.043 | 0.032 | 0.023 | 0.041 |
| PRGBP | -0.434 | -0.440 | -0.428 | -0.348 | -0.382 | -0.309 | -0.313 | -0.327 | -0.299 | -0.268 | -0.291 | -0.245 |
| PRGDM | -0.177 | -0.186 | -0.169 | -0.192 | -0.243 | -0.151 | -0.178 | -0.196 | -0.161 | -0.180 | -0.199 | -0.162 |
| SEXINF | -0.041 | -0.049 | -0.032 | -0.003 | -0.025 | 0.018 | -0.060 | -0.077 | -0.043 | -0.100 | -0.121 | -0.079 |

Legend: Intercepts are path coefficients from causal factors to duration of gestation (OBGEST), from Bayesian estimation, together with their lower and upper 95% credibility intervals. Coefficients for variables that increase OBGEST in ethnic minorities (95%CIs do not overlap those of nHEA) are in green; those that decrease OBGEST are in red. Key: as in Tables S7a-b.

Table S7i: Variances of continuous variables for each of ethnic groupings

| Ethnicity: | Non-Hispanic European American | | | African-American | | | Hispanic | | | Other (mostly Asian) | | |
|---|---|---|---|---|---|---|---|---|---|---|---|---|
| Variable | Intercept | low 95%CI | high 95%CI | Intercept | low 95%CI | high 95%CI | Intercept | low 95%CI | high 95%CI | Intercept | low 95%CI | high 95%CI |
| FAGE | 1.028 | 1.021 | 1.035 | 1.425 | 1.404 | 1.447 | 1.19 | 1.173 | 1.207 | 1.071 | 1.058 | 1.085 |
| MAGE | 0.591 | 0.586 | 0.596 | 0.598 | 0.583 | 0.614 | 0.666 | 0.654 | 0.677 | 0.641 | 0.631 | 0.651 |
| MHT | 0.443 | 0.441 | 0.445 | 0.461 | 0.456 | 0.466 | 0.379 | 0.376 | 0.383 | 0.339 | 0.335 | 0.343 |
| BMI | 0.907 | 0.898 | 0.916 | 1.049 | 1.007 | 1.087 | 0.969 | 0.958 | 0.980 | 0.623 | 0.609 | 0.635 |
| PRECAREM | 1.275 | 1.27 | 1.281 | 2.206 | 2.161 | 2.244 | 2.278 | 2.257 | 2.299 | 1.949 | 1.921 | 1.975 |
| OBGEST | 1.423 | 1.416 | 1.431 | 1.438 | 1.368 | 1.483 | 1.425 | 1.409 | 1.440 | 1.381 | 1.359 | 1.402 |
| BWT | 0.186 | 0.185 | 0.187 | 0.180 | 0.169 | 0.188 | 0.181 | 0.179 | 0.183 | 0.173 | 0.171 | 0.175 |

Legend: Intercepts are variances from Bayesian estimation, together with their lower and upper 95% credibility intervals. Variances that are greater in ethnic minorities (95%CIs do not overlap those of nHEA) are in red; those that are smaller are in green. Key: FAGE = paternal age; MAGE = maternal age; MHT = maternal height; BMI = pre-pregnancy maternal BMI; PRECAREM = month of gestation when pre-natal care began; OBGEST = duration of gestation (obstetric estimate); BWT = birthweight.

Table S7j: Miscellaneous effects of different demographic variables on each other

| Ethnicity | | Non-Hispanic European American | | | African-American | | | Hispanic | | | Other (mostly Asian) | | |
|---|---|---|---|---|---|---|---|---|---|---|---|---|---|
| Cause | effect | Intercept | low 95%CI | high 95%CI | Intercept | low 95%CI | high 95%CI | Intercept | low 95%CI | high 95%CI | Intercept | low 95%CI | high 95%CI |
| FAGE | FEDUC | 0.003 | -0.006 | 0.011 | 0.071 | 0.059 | 0.082 | -0.063 | -0.079 | -0.048 | 0.012 | -0.002 | 0.025 |
| CIG0 | MAGE | 0.011 | 0.006 | 0.017 | 0.019 | 0.002 | 0.036 | -0.027 | -0.047 | -0.007 | 0.021 | -0.004 | 0.046 |
| CIG0 | MEDUC | -0.120 | -0.134 | -0.107 | -0.113 | -0.146 | -0.079 | -0.069 | -0.099 | -0.039 | -0.001 | -0.055 | 0.053 |
| MEDUC | BMI | 0.126 | 0.114 | 0.138 | 0.226 | 0.187 | 0.268 | 0.084 | 0.070 | 0.098 | 0.088 | 0.064 | 0.114 |
| MAGE | BMI | 0.216 | 0.207 | 0.226 | 0.354 | 0.311 | 0.398 | 0.199 | 0.185 | 0.213 | 0.047 | 0.032 | 0.063 |
| CIG0 | BMI | -0.035 | -0.043 | -0.026 | 0.040 | 0.014 | 0.067 | 0.023 | -0.003 | 0.050 | 0.059 | 0.027 | 0.090 |

Legend: Intercepts are path coefficients from causal factors to effects, from Bayesian estimation, together with their lower and upper 95% credibility intervals. Coefficients that are higher in ethnic minorities (95%CIs do not overlap those of nHEA) are in red; those that are lower are in green. Key: as in Table S7a-b..

Figure S7k: correlations between demographic factors

| Ethnicity: | | Non-Hispanic European American | | | African-American | | | Hispanic | | | Other (mostly Asian) | | |
|---|---|---|---|---|---|---|---|---|---|---|---|---|---|
| X1 ↔ X2 | | Intercept | lo 95%CI | hi 95%CI | Intercept | lo 95%CI | hi 95%CI | Intercept | lo 95%CI | hi 95%CI | Intercept | lo 95%CI | hi 95%CI |
| FAGE | MAGE | 0.526 | 0.521 | 0.531 | 0.599 | 0.584 | 0.614 | 0.573 | 0.561 | 0.585 | 0.511 | 0.501 | 0.52 |
| FEDUC | MEDUC | 0.200 | 0.188 | 0.214 | 0.327 | 0.308 | 0.345 | 0.356 | 0.342 | 0.371 | 0.437 | 0.408 | 0.464 |
| MEDUC | MAGE | 0.072 | 0.067 | 0.078 | 0.058 | 0.046 | 0.071 | 0.085 | 0.076 | 0.094 | 0.089 | 0.078 | 0.100 |
| PAYER | WIC | -0.526 | -0.536 | -0.515 | -0.316 | -0.343 | -0.288 | -0.503 | -0.520 | -0.485 | -0.410 | -0.468 | -0.357 |
| PAYER | BMI | 0.175 | 0.167 | 0.184 | 0.189 | 0.162 | 0.218 | 0.098 | 0.083 | 0.113 | 0.145 | 0.118 | 0.174 |
| PAYER | CIG0 | -0.118 | -0.129 | -0.106 | -0.103 | -0.142 | -0.064 | -0.107 | -0.153 | -0.062 | 0.138 | 0.053 | 0.225 |
| WIC | BMI | -0.073 | -0.087 | -0.060 | -0.187 | -0.276 | -0.112 | -0.013 | -0.027 | 0.001 | -0.116 | -0.186 | -0.066 |
| WIC | CIG0 | 0.181 | 0.168 | 0.194 | 0.105 | 0.061 | 0.150 | 0.024 | -0.012 | 0.060 | -0.101 | -0.225 | 0.006 |
| PRGBP | PNC | -0.023 | -0.029 | -0.017 | 0.030 | 0.002 | 0.060 | -0.003 | -0.022 | 0.016 | 0.003 | -0.027 | 0.033 |
| PRGDM | PNC | 0.003 | -0.005 | 0.011 | 0.004 | -0.038 | 0.040 | -0.006 | -0.028 | 0.016 | -0.037 | -0.059 | -0.016 |
| PRGBP | PRGDM | 0.065 | 0.054 | 0.076 | 0.141 | 0.109 | 0.176 | 0.143 | 0.118 | 0.167 | 0.087 | 0.056 | 0.117 |
| OBGEST | BWT | 0.234 | 0.232 | 0.236 | 0.239 | 0.214 | 0.258 | 0.228 | 0.224 | 0.232 | 0.231 | 0.225 | 0.237 |

Legend: Intercepts are means of path coefficients from causal factors to duration of gestation (OBGEST), from Bayesian estimation, together with their lower and upper 95% credibility intervals. Correlations that are absolutely smaller in ethnic minorities (95%CIs do not overlap those of nHEA) are in green; those that are absolutely larger are in red. Key: as in Tables S7a-b.

Table S7l: Thresholds of ordinal variables for each of the main ethnic groupings

| Ethnicity: | Non-Hispanic European American | | | African-American | | | Hispanic | | | Other (mostly Asian) | | |
|---|---|---|---|---|---|---|---|---|---|---|---|---|
| Variable | Intercept | low 95%CI | high 95%CI | Intercept | low 95%CI | high 95%CI | Intercept | low 95%CI | high 95%CI | Intercept | low 95%CI | high 95%CI |
| FEDUC-1 | -2.636 | -2.667 | -2.605 | -1.717 | -1.744 | -1.692 | -1.019 | -1.037 | -1.002 | -2.521 | -2.631 | -2.428 |
| FEDUC-2 | -0.877 | -0.889 | -0.865 | 0.012 | -0.001 | 0.026 | 0.435 | 0.416 | 0.454 | -1.477 | -1.545 | -1.420 |
| FEDUC-3 | -0.101 | -0.107 | -0.094 | 0.853 | 0.833 | 0.875 | 1.232 | 1.201 | 1.263 | -0.976 | -1.024 | -0.935 |
| FEDUC-4 | 0.248 | 0.241 | 0.254 | 1.14 | 1.116 | 1.165 | 1.588 | 1.551 | 1.625 | -0.739 | -0.778 | -0.705 |
| FEDUC-5 | 2.585 | 2.555 | 2.615 | 3.026 | 2.972 | 3.084 | 3.363 | 3.288 | 3.435 | 1.796 | 1.737 | 1.868 |
| MEDUC-1 | -3.217 | -3.259 | -3.179 | -2.162 | -2.212 | -2.114 | -1.412 | -1.434 | -1.391 | -2.766 | -2.84 | -2.696 |
| MEDUC-2 | -1.513 | -1.534 | -1.495 | -0.376 | -0.394 | -0.358 | 0.008 | -0.004 | 0.019 | -1.699 | -1.745 | -1.655 |
| MEDUC-3 | -0.608 | -0.618 | -0.598 | 0.677 | 0.657 | 0.698 | 0.870 | 0.855 | 0.886 | -1.161 | -1.194 | -1.129 |
| MEDUC-4 | -0.152 | -0.159 | -0.145 | 1.054 | 1.027 | 1.081 | 1.247 | 1.227 | 1.267 | -0.876 | -0.903 | -0.85 |
| MEDUC-5 | 2.879 | 2.844 | 2.917 | 3.508 | 3.433 | 3.591 | 3.388 | 3.337 | 3.441 | 2.072 | 2.018 | 2.126 |
| CIG0 | 1.794 | 1.783 | 1.806 | 2.054 | 2.029 | 2.08 | 2.376 | 2.351 | 2.402 | 2.916 | 2.822 | 3.012 |
| PAYER | -1.181 | -1.195 | -1.167 | 0.303 | 0.287 | 0.32 | 0.25 | 0.231 | 0.27 | -1.288 | -1.386 | -1.199 |
| WIC | 1.397 | 1.374 | 1.426 | -0.049 | -0.063 | -0.034 | -0.022 | -0.032 | -0.012 | 1.563 | 1.366 | 1.83 |
| PRGBP | 1.377 | 1.367 | 1.388 | 1.365 | 1.343 | 1.387 | 1.549 | 1.526 | 1.573 | 1.806 | 1.748 | 1.874 |
| PRGDM | 1.828 | 1.815 | 1.841 | 1.935 | 1.891 | 1.991 | 1.812 | 1.785 | 1.84 | 1.400 | 1.367 | 1.434 |

Values are unstandardised mean values and their 95% credibility intervals.  values for the thresholds are probits. Note also that for binary variables, Mplus reports higher thresholds for rarer events. I have colour-coded values for minority ethnicities where 95% CIs do not overlap non-Hispanic European Americans' (NHEA): red = (notionally) worse than nHEA; green = (notionally) better than nHEA

**Separate analyses of the "realistic" model for each ethnicity**

Tables S7a-d show the direct effects of demographic factors, except ethnicity, separately for each ethnic group. These analyses parallel the 'realistic' model in the main report, except that they analysed each ethnic group separately and omitted all direct effects of ethnicity. Hence, comparisons between ethnicities in these tables allow assessment of heterogeneity of interactive effects of demographic factors with ethnicity. I mention briefly here only the major differences, comparing each minority ethnicity with nHEA:-

1) The loadings of demographic variables on the factor of social deprivation differed substantially between nHEA and ethnic-minority births (Table S7a). Specifically:-
a) Age loaded more strongly on deprivation in AA and Hispanic fathers
b) Both paternal and maternal education loaded less on deprivation in all ethnic-minority parents. This may reflect discrimination in employment.
c) Maternal height loaded less on deprivation in AA and Other-ethnicity mothers, but more in Hispanic mothers.
d) Smoking loaded less on deprivation in AA and Hispanic mothers.
e) BMI loaded less on deprivation in Hispanic and Other-ethnicity mothers

2) Deprivation had much worse direct effects on birth outcomes in AA parents, but had mixed effects in Hispanic and Other-ethnicity parents (Table S7b).

3) More educated mothers of all ethnic-minorities more often participated in WIC (Table S7c).

4) Older or more educated mothers of all ethnic minorities began pre-natal care earlier (Table S7d).

5) Effects of demographic factors on pregnancy hypertension and gestational diabetes showed relatively little heterogeneity between ethnic groups (Tables S7e-f).

6) Effects of demographic factors on birthweight showed substantial heterogeneity (Table S7g).
a) Birthweights were less for AA mothers who paid privately, smoked, were older or more educated
b) Boy babies of Hispanic or Other ethnicity were relatively lighter.
c) Surprisingly, Other-ethnicity mothers who smoked had heavier babies
d) Participation in WIC increased birthweight in all ethnic minorities.

7) Effects of demographic factors on duration of gestation showed substantial heterogeneity (Table S7h).
a) later initiation of pre-natal care lengthened gestation *less* for all ethnic minority mothers than for nHEA mothers.
b) Participation in WIC lengthened gestation in all ethnic minorities.
c) Pregnancy hypertension shortened gestation in all ethnic minorities *less* than in nHEA mothers.
d) Private payment, smoking, more education and older maternal age all shortened gestation in AA mothers.

8) Correlations between demographic factors
a) Private payment and participation in WIC showed weaker inverse correlations in all ethnic minorities, compared with nHEA.
b) Maternal and paternal education correlated more strongly in all ethnic minorities than in nHEA.

In summary, there is substantial heterogeneity between different ethnicities in all aspects of the realistic model. This may partly explain why the model's imperfect fit (see Main Report).

**Supplementary Discussion**

*Exclusions*

I excluded *a priori* births to mothers who had prepregnancy hypertension or diabetes, or extreme physical characteristics, or of babies born prematurely or at very low birthweight. These cases had worse outcomes. So, their exclusion probably diminished effect sizes of variables in the study, for the population as a whole. On the other hand, if I had included these cases, then could distort the results because poor outcomes in the small numbers of these cases could exert undue leverage in the analysis. Therefore, I chose to omit these atypical cases in order to ensure that the present results are more likely to be relevant to most primigravidae. Further studies could extrapolate the realistic model (see Main Report) to determine how well it can predict the outcomes of excluded cases.

*Missing data*

There is clear evidence that missing data are "missing not at random" (MNAR). That is, the causes of missingness – particularly paternal ethnicity and education – also cause obstetric outcomes (see Tables 4-5 in Main Report). Tables S2-S4 indicate that (a) cases with partial missing data have worse outcomes, (b) maternal ethnicity may cause missingness, and (c) patterns of missingness are very different for mothers of minority ethnicities, compared with nHEA mothers. Hence, overall, the Main Report's analysis of cases with complete data most likely *under*-estimates adverse effects of minority ethnicity. Since parental minority ethnicity also strongly predicts deprivation, it is also probable that the Main Reports model *under*-estimates effects of deprivation on outcomes.

It is uncertain if data imputation would improve estimates of ethnicity and deprivation on outcomes. Random forest procedures for imputation can cope with non-linear causal effects and complex interactions,[16] but may still result in severely biased estimates of missing values.[17] Potentially, Mplus analyses can themselves impute missing data in the context of the model.[6] However, the accuracy of such imputation would depend on model fit – which here is imperfect – and on model validity, which is uncertain (see below). In the present context, imputation should ideally incorporate geopolitical and financial factors that may determine missingness.

*The 'no-racism' model*

I do not describe the 'no-racism' model in detail, for three reasons. First, the realistic model can fit the data, but the 'no-racism' model cannot. Second, the 'no-racism' model appears implausible. For example, it implies that smoking *causes* younger maternal age, whereas the realistic model views both smoking and younger maternal age as *effects* of deprivation. Third, the realistic model's results parallel external causal evidence – e.g. that smoking increases maternal age (due to smoking's toxicity – see Main Report), *contra* the 'no-racism' model's result (see Mplus outputs). Therefore, I present the 'no-racism' model only highlight its inadequacies, in relation the the realistic model.

*Model interpretation*

The model presents an "open theory" of the data-generating processes for obstetric outcomes.[18] The fact that the model can fit a 1% sample of the data indicates that the model's paths can bear interpretation as causal effects. Appropriate combinations of the model's direct effects (see Tables 3-5 in Main Report) can quantify indirect effects, to test how far intermediate variables may mediate total causal effects. In general, deprivation mediated many – but not all – effects of minority ethnicity.

The underlying rationale for the present study is that linear structural equation models can be a "useful microscope" for causal analysis.[19] However, causal interpretations depend on unmet "cross-world counterfactual assumptions".[20] The presence of variables (measured or unmeasured) that result from a 'treatment' and in turn cause both mediators and outcomes is guaranteed to violate these assumptions.[21] The present model includes variables of this kind. For, example,  if we regard maternal ethnicity as a 'treatment', this causes social deprivation, which in turn can cause both mediators – e.g. maternal education, height, or smoking – and outcomes, e.g. birthweight. Unknown variables may also violate the cross-world assumption. For example, rurality may determine proximity to a teaching hospital, which in turn may influence both the nature of care and outcomes.[22] It is possible that observational data can never fulfil the cross-world assumption in obstetric settings. However, the present study may help to highlight questions that merit more detailed study, using randomised controlled trials.

*Statistical considerations*

Model fit

Assessing model fit is a general problem.[10] With large N, $\chi^2$ may be highly sensitive for detecting poor fit, but other fit indices may be insensitive. My analysis of a 1% sample (7908 cases) here may achieve an adequate balance between sensitivity and misspecification – it has 42 cases per path coefficient, but fits according to the posterior predictive $\chi^2$ (95% CI = -26.140 – 710.705, posterior predictive probability =0.132).

The Main Report discusses possible reasons for its model's imperfect fit. Additional remediable causes are: (i) the model used the probit link for categorical variables (which is intolerant of outliers); (ii) the model used identity links for continuous variables (e.g. age, BMI, height), even though appropriate transformations can improve model fit, overall.[24]

Effect sizes

The realistic model's causal effects are generally small. Two possible reasons for this are (1) the study excluded cases with missing data, but these cases showed worse outcomes (for available data). Hence, it is likely that the model under-estimated causal effects. (2) The model structure allows "false witness" effects that render causal paths non-identified and could dilute causal estimates.[21] Nevertheless, (a) even small effects in the study surpass the strict criterion of the 't' approximation to the Bayes Factor that I adopted here[25] – which indicates that the data show very strong evidence for them (b) small effects are often more realistic[26] and together can add up (although many indirect effects did not reinforce each other but cancelled each other's effects); (c) the omission of obstetric interventions from the model may partly obscure some causal effects on birth outcomes (see below). In spite of these limitations, the present model may help to guide clinicians' reasoning about causes and treatments of some adverse outcomes.

Model incompleteness – effects of obstetric interventions

A further limitation of the present study is that it omits effects of obstetric management on outcomes. However, it is unlikely that this omission obscures these causal effects completely, because active obstetric management (e.g. induction, or Caesarean section) is often a response to pregnancy complications – gestational hypertension and diabetes – that are present in the model.

Hence, active obstetric management does not obscure causal paths to these *pregnancy* complications, but may distort their apparent effects on *birth* outcomes – birthweight and duration of gestation. Elective delivery, for non-obstetric reasons, may also affect birth outcomes – and the model does not account for these effects. It is likely that elective delivery is more frequent when mothers have private insurance or pay directly, but emergency delivery is more likely in mothers who suffer more deprivation. Further studies should extend the present model to include obstetric management and ascertain its effects on outcomes more fully.

21 Avin C, Shpitser I, Pearl J. Identifiability of Path-Specific Effects. .

22 Vinekar K, Qasba N, Reiser H, *et al.* Segregation by Payer in Obstetrics and Gynecology Residency Ambulatory Care Sites. *Med Educ*.

23 Wong MS, Steers WN, Hoggatt KJ, Ziaeian B, Washington DL. Relationship of neighborhood social determinants of health on racial/ethnic mortality disparities in US veterans-Mediation and moderating effects. *Health Serv Res* 2020; **55 Suppl 2**: 851–62.

24 Hothorn T, Zeileis A. Predictive Distribution Modeling Using Transformation Forests. *J Comput Graph Stat* 2021; **30**: 1181–96.

25 Raftery AE. Bayesian model selection in social research. *Sociol Methodol* 1995; **25**: 111–63.

26 Götz FM, Gosling SD, Rentfrow PJ. Small Effects: The Indispensable Foundation for a Cumulative Psychological Science. ; : 11.
